# White Matter Microstructure Alterations in Early Psychosis and Schizophrenia

**DOI:** 10.1101/2024.02.01.24301979

**Authors:** Tommaso Pavan, Yasser Alemán-Gómez, Raoul Jenni, Pascal Steullet, Zoé Schilliger, Daniella Dwir, Martine Cleusix, Luis Alameda, Kim Q. Do, Philippe Conus, Paul Klauser, Patric Hagmann, Ileana Jelescu

**Affiliations:** Department of Radiology, Lausanne University Hospital (CHUV) and University of Lausanne (UNIL); Center for Psychiatric Neuroscience, Department of Psychiatry, Lausanne University Hospital and the University of Lausanne, Lausanne Switzerland; Service of General Psychiatry, Treatment and Early Intervention in Psychosis Program. Lausanne University Hospital (CHUV), Lausanne, Switzerland; Department of Psychosis Studies, Institute of Psychiatry, Psychology and Neuroscience. King’s College of London, London, UK; Centro Investigacion Biomedica en Red de Salud Mental (CIBERSAM); Instituto de Biomedicina de Sevilla (IBIS), Hospital Universitario Virgen del Rocio, Departamento de Psiquiatria, Universidad de Sevilla, Sevilla, Spain; Service of Child and Adolescent Psychiatry, Department of Psychiatry, Lausanne University Hospital and the University of Lausanne, Lausanne, Switzerland

## Abstract

Studies on schizophrenia feature diffusion magnetic resonance imaging (dMRI) to investigate white matter (WM) anomalies. The heterogeneity in the possible interpretations of typical Diffusion Tensor Imaging (DTI) metrics highlights the importance of increasing their specificity. Here, we characterize WM pathology in early psychosis (EP) and schizophrenia (SZ) with increased specificity using advanced dMRI: Diffusion Kurtosis Imaging and the biophysical model White Matter Tract Integrity – Watson (WMTI-W). This enables us to better characterize WM abnormalities, while preserving good sensitivity to group differences, and relate them to the current literature (ENIGMA-schizophrenia), patient’s clinical characteristics and symptomatology. dMRI-derived microstructure features were extracted from all of WM and from individual regions of interest in 275 individuals. 93 subjects diagnosed with EP and 47 with SZ were compared respectively to 135 age-range matched healthy controls (HC). WM DTI diffusivities were higher, while kurtosis was lower in EP vs HC and in SZ vs HC. Differences were more widespread in EP than SZ. The regional alterations found in our cohort matched the DTI patterns found in ENIGMA-schizophrenia. WMTI-W model parameters indicate that the WM alterations in patients come primarily from the extra-axonal compartment, consistent with abnormal myelin integrity in the disease pathology. The direct link between WM alterations and symptomatology is, however, limited.

## Introduction

Psychosis is a psychiatric disorder with heavy implications for the affected individuals, their families, and society^1^. However, the disorder etiology remains only partially understood and is considered multifactorial, involving a complex interplay of genetic, environmental, and neurobiological factors^2,3^. Many clinical studies investigating the psychosis spectrum^4–9^ reported pathological white matter (WM) as a common feature of the disease. Postmortem studies further substantiate the WM involvement found in first-episode psychosis participants^3^ and in SZ, reporting splitting and decompacting of myelin sheaths together with dystrophic oligodendroglia^10–13^ and microglia^14^.

Most in vivo studies of WM abnormalities in psychosis use diffusion magnetic resonance imaging (dMRI), an MRI technique that relies on the random motion of water molecules to explore the cellular environment and thus infer the microstructural properties of the underlying biological tissue^15^. The most popular dMRI technique in clinical research is Diffusion Tensor Imaging (DTI) and its derived scalar metrics: fractional anisotropy (FA), mean/axial/radial diffusivities (MD/AD/RD). DTI is a so-called *signal representation*^16^, which means it makes no assumption about the underlying tissue and reports on the apparent diffusivity in the voxel, in any given direction of space^17^.

In schizophrenia (SZ) and early psychosis (EP), the most frequently reported and accepted dMRI patterns are reduced FA and increased RD and MD^5,8,18–21^, consistent with a loss of WM integrity and reduced diffusion barriers.

ENIGMA-schizophrenia, the largest consortium to date to coordinate an effort across multiple sites to map the WM alterations in SZ, found widespread changes with reproducible patterns across the WM in their meta-analysis^8,22^. The most affected regions were reported to be the anterior corona radiata (ACR) and the corpus callosum^8^ (CC). However, the largest effect size between patients and controls was typically found in the average WM skeleton, which seemed to drive the difference also across more specific regions of interest^8^ (ROI). In early-onset psychosis, the superior longitudinal fasciculus (SLF), posterior thalamic radiation (PTR) and superior fronto-occipital fasciculus (SFO) were also reported to be affected^6^. According to the developmental interpretation of schizophrenia, alterations originate during adolescence and remain throughout the lifespan^23–26^. It is therefore expected for EP to show agreement with chronic SZ in the spatial distribution of WM alterations, albeit with reduced magnitude, as shown in Barth et al^6^.

However, while DTI metrics are sensitive to WM changes, they only provide a coarse depiction of microstructure. A first step towards improving the microstructure characterization is to estimate the tissue diffusion properties more thoroughly, e.g. using Diffusion Kurtosis Imaging^27,28^ (DKI), an extension of DTI that is estimated from stronger diffusion-weighting. DKI provides complementary information about tissue heterogeneity by going beyond the Gaussian DTI approximation. DKI has been able to detect widespread WM abnormality^29^ in regions with complex fiber arrangement^30^, subtle abnormalities in subjects at high risk for psychosis^31^ and microstructural connectivity patterns associated to processing speed deficits in SZ^32^. Nevertheless, DTI and DKI metrics are only *sensitive* to features of the tissue microstructure and their changes can be the consequence of several possible pathological mechanisms (**Table 1)**. To gain specificity, WM *biophysical models* of diffusion are used^16^, that capture the diffusion behavior in the underlying tissue with a mathematical model based on *a-priori* knowledge of the tissue structure. Models previously used to characterize EP and SZ are free-water imaging^33^ (FWI) and Neurite Orientation Dispersion and Density Imaging^34^ (NODDI). Most FWI studies reported a global increase in free-water in SZ^35^ and across lifespan^36^. NODDI detected decreased neurite density and increased orientation dispersion index in first-episode^37,38^ and SZ^39^. However, both models display limitations in terms of *ad hoc* simplifying assumptions and fit constraints (e.g. in NODDI, all three compartment diffusivities are fixed^34,40^), strongly limiting the interpretability and validity^40^ of the microstructure parameters estimated from the data^16,41^.

**Table 1:**
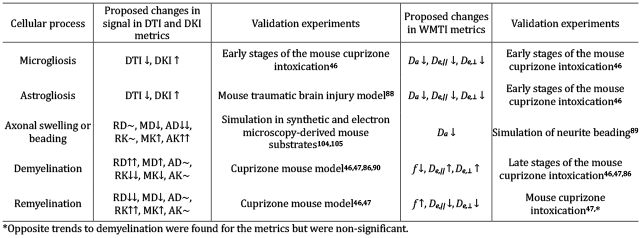
Overview of the effect of various pathological processes on diffusion metrics, adapted from ^44^. Astro- and microgliosis reduce water diffusivity and increase kurtosis via increased cellularity. Demyelination on the contrary translates to increased diffusivities and reduced kurtosis. When both are concomitant, they have competing effects. ↑: increase, ↓: decrease, ∼: unchanged, DTI, DKI diffusion tensor and kurtosis imaging, RD, RK, MD, MK, AD, AK: Radial, Mean, Axial Diffusivity or Kurtosis, *f* : axonal water fraction, *D*_*a*_: intra-axonal diffusivity, *D*_*e,//*_, *D*_*e*,⊥_:extra-axonal parallel and radial diffusivities.

To overcome these limitations, we used White Matter Tract Integrity-Watson^42,43^ (WMTI-W), which enables the estimation of intra-(*D*_*a*_) and extra-axonal (*D*_e,∥_, *D*_e,⊥_) specific diffusivities that are excellent proxies for intra-axonal injury, inflammation and abnormal myelin integrity respectively^16,43,44^, in addition to axonal density (*f*) and orientation dispersion (*c*_*2*_). This unconstrained two-compartment model of WM has been recently used to characterize WM in a variety of pathological mechanisms (**Table 1**), patient populations^45^ and animal models of disease^46,47^, ranging from Alzheimer’s disease^48–50^ to traumatic brain injury^51^, but, to our knowledge, never in schizophrenia.

This work is articulated around two main hypotheses:

i. our Lausanne Psychosis cohort (LSP) will exhibit good consistency in WM microstructure changes characterized by DTI and DKI with the current state of the literature as reported by ENIGMA^8^.
ii. WMTI-W metrics allow the characterization of WM pathology in EP and SZ with increased specificity and limited penalty in sensitivity, as compared to the ‘omnibus’ DTI/DKI metrics. In light of previous post-mortem ultrastructural findings of myelinated fiber pathology (myelin sheath splitting with inclusions of vacuoles, small-axons atrophy, dystrophic oligodendroglia^10–13^) and dMRI literature (widespread increased MD, RD and reduced FA^8^), we expect that our biophysical findings will be mainly reflected as increased diffusivity of the extra-axonal space and as reduced axonal density.

In an exploratory fashion, we also related WM pathology as characterized by diffusion MRI to commonly used clinical measures and characteristics, such as age of psychosis onset, illness duration, medication, and symptomatology.

## Methods

### Participants

The study was approved by the local Ethics Committee of the Canton of Vaud (CER-VD, Switzerland). Data was collected from 135 healthy controls (HC), 93 subjects with EP and 47 with SZ (**Table 2**). Participants with a diagnosis of EP (within 5 years after a first psychotic episode as defined by the CAARMS^52^) or SZ (DSM-IV diagnosis of schizophrenia or schizoaffective disorder) were recruited from the Lausanne University Hospital. Exclusion criteria were psychosis related to intoxication, organic brain disease, IQ*<*70, alcoholism, drug abuse, major somatic disease, or current organic brain damage. Duration of illness, age at psychosis onset and medication were registered. HC were recruited from the same sociodemographic area of the clinical groups and were excluded if they had a first-degree family member who suffered from psychosis or prodromal symptoms, or if they reported current or past antipsychotic treatment. The HC group was further subdivided into younger (HC_Young_, n=130, age=26.8±6.8) and older (HC_Old_, n=84, age=31.9±8.1) to better match the age ranges of the two clinical groups (EP=24.7±5.5; SZ=38.1±9.4 years). Diagnoses are reported in **Table S1**.

**Table 2:**
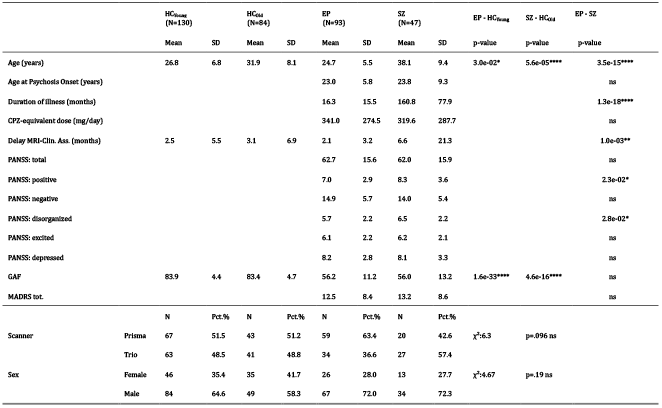
Cohort demographics. HC amount to 135 Individuals, 79 HC were shared between HC_Young_ and HC_Old_. P-values refer to Wilcoxon’s tests between clinical and HC groups. *χ*^2^ test is computed for the scanner and sex contingency table. CPZ: chlorpromazine, Delay MRI-Clin. Ass.: difference in months between MRI scan and clinical assessment, GAF: Global Assessment of Functioning, PANSS: Positive and Negative Syndrome Scale, MADRS: Montgomery-Asberg Depression Rating Scale.

### MRI acquisition

MRI data was acquired on two different 3-Tesla systems (Magnetom TrioTim and PRISMA, Siemens Healthineers, Erlangen, Germany), each equipped with a 32-channel head coil. A 1-mm isotropic T1-weighted image was acquired for anatomical reference. Whole-brain diffusion-weighted images (DWI) were acquired using diffusion spectrum imaging (DSI) scheme across 15 b-values, ranging from 0 to 8000 s/mm^2^, voxel size of 2.2 × 2.2 × 3 mm^3^. Further information can be found in the ‘Acquisition details’ section of the Supplementary Material.

### Image preprocessing

The T1-weighted images were bias field corrected^53^ and skull-stripped via nonlinear registration to the MNI-152 template using Advanced Normalization Tools^54^ (ANTs). The diffusion preprocessing pipeline included MP-PCA denoising, Gibbs ringing-, EPI-, eddy current and motion corrections, following most recent guidelines^55^ - see Supplementary Material for preprocessing details.

### Microstructure estimation

For DKI and WMTI-W estimation, the diffusion dataset was truncated^27^ at b≤2500 s/mm^2^. DKI was fit voxel-wise^56^ using Matlab, from which seven parameter maps were derived. Four from DTI: RD, MD, AD and FA, and three from DKI: radial, mean, axial kurtosis (RK, MK, AK). Then, WMTI-W model parameters were estimated voxel-wise from the DTI and DKI parameters, using an in-house Python script, yielding other five parameter maps: axonal density *f*, intra-axonal diffusivity *D*_a_, extra-axonal parallel and perpendicular diffusivities *D*_e,∥_, *D*_e,⊥_ and axon orientation alignment *c*_2_. The WM characterization thus relied on 12 microstructure metrics. Code is available here: https://github.com/tmspvn/python-WMTI-watson.

### ROI analysis

Individual FA maps were non-linearly registered to the Johns Hopkins University FA template^57^ (JHU) using ANTs, and the WM region-of-interest (ROI) atlas labels were mapped back to individual space. The mean value of each microstructure metric was computed for each ROI, as well as in the whole-WM mask defined as the collection of all JHU ROIs (from here on referred to as ‘WM core’). It should be noted that our ROI definition slightly differs from the ENIGMA ROI definition (JHU atlas intersected with the WM skeleton from TBSS analysis^58^). Our choice was guided by several factors. First, this enabled us to perform our analysis in native space instead of standard space, which reduces mis-alignment problems^59,60^ and completely avoids interpolation and non-linear deformations of quantitative diffusion metrics to standard space. Second, by averaging full-size ROIs instead of their intersection with the WM skeleton, we are able to study the whole volume of the WM tract instead of only the contribution of its core as defined by the maximum FA value^58^. This allows us to better account also for potential alterations at the cross-sectional boundaries/edges of the tract. Finally, our ROI definition enables us to report WM differences also in the cerebellum, an important region in schizophrenia^61^, that are not reported in ENIGMA.

### Psychiatric scales

Psychiatric tests included the Global Assessment of Functioning^62^ scale (GAF), the Positive and Negative Syndrome Scale^63^ (PANSS), and the Montgomery-Asberg Depression Rating Scale^64^ (MADRS). For the PANSS, items were categorized using the Wallwork/Fortgang five-factor model^65^. PANSS data was not available for 5 EP and 1 SZ subjects, MADRS was not assessed in 6 EP and 16 SZ subjects.

### Statistical analysis

Before any statistical analysis, all the microstructure parameter estimates in the WM core and individual JHU ROIs were harmonized for scanner type via ComBat harmonization^66^ that was proven efficient at correcting scanner effects in the same cohort^67^. Distributions for each metric, ROI and group were tested for normality using the Shapiro–Wilk test and for homogeneity of variance using the Levene’s Test. The statistical test used for group comparisons was chosen based on distribution characteristics, resulting in microstructure metrics for the WM core being tested via non-parametric *Wilcoxon* signed-rank test, suitable for non-normal but homogeneous variance distributions. At the ROI level, microstructure metrics between groups were compared using the non-parametric *Brunner-Munzel*^68,69^ test, suitable for distributions with unequal variances. In all comparisons, the estimates were controlled for sex and quadratic age to properly account for age differences between groups (see ‘Age correction’ section in Supplementary Material, and **Fig. S1**). False-Discovery Rate (FDR) correction was applied to control for false positives. In total, 24 tests (12 metrics × 2 group pairs) were conducted for group differences between metrics in the WM core, and 1200 tests for ROI-specific group differences (12 metrics × 50 JHU ROIs × 2 group pairs). Finally, we tested what ROI differences would survive the inclusion of the core WM as a covariate.

Effect sizes were estimated using *Cohen’s d*. The dice coefficient was used to quantify the similarity in ROI alterations across the brain, indicating the proportion (between 0 and 1) of significant alterations that EP and SZ groups have in common. Agreement between effect sizes previously reported in the ENIGMA ROIs and the LSP cohort, as well as between EP vs SZ within the LSP cohort were evaluated via correlations.

Associations of the microstructure with clinical characteristics and with psychiatric scales was done via regression analysis of the 12 dMRI metrics in the WM core with age at first psychosis, duration of illness, chlorpromazine-equivalent dose (CPZ-equivalent) and psychiatric symptoms scales were estimated in R^70^. Each association was corrected for quadratic age and sex, and p-values were FDR corrected. Between-group differences in aging trajectories were similarly estimated, but pooling HC vs clinical participants (EP+SZ). Symptoms scale correlations were analyzed via hierarchical clustering and FDR corrected, after excluding participants with a delay between clinical assessment and MRI scan larger than 45 days, leaving 58 EP (mean delay 0.43±0.60 months) and 33 SZ (0.58±0.42 months).

## Results

### Demographics

**Table 2** collects cohort demographics. Age was significantly different between clinical participants and controls (i.e., EP vs HC_Young_, p=0.03) and SZ vs HC_Old_, p*<*.0001), hence the need for quadratic age correction. Illness duration was longer in SZ than in EP group (p*<*.0001).

### Microstructure imaging estimates in the white matter core

Compared to HC_Young_, EP showed significantly higher DTI diffusivities and lower FA (**Fig. 1A-D**; RD, MD, FA: p<.0001, AD: p=.014), as well as lower kurtosis (**Fig. 1E-G**; RK: p=.0017, MK: p=.0029; AK: p=.035). Axonal water fraction, *f*, and alignment, *c*_*2*_, in EP were also significantly lower than in HC_Young_ (**Fig. 1H, L; *f***: p=.0053 *c*_2_: p=.0010), while extra-axonal diffusivities, *D*_*e,∥*_ and *D*_*e*,⊥_, were higher (**Fig. 1J, K**; *D*_*e,∥*_: p=.0010, *D*_*e*,⊥_: p<.0001). No significant differences were found in terms of intra-axonal diffusivity *D*_*a*_ (**Fig. 1I**). Effect sizes were medium to high (mean |*d*|=.46) with the highest being RD (*r*=.64), followed by FA (*r*=-.62), MD (*r*=.60), and D_e,⊥_ (*r*=.59) (**Table S2**).

**Figure 1.**
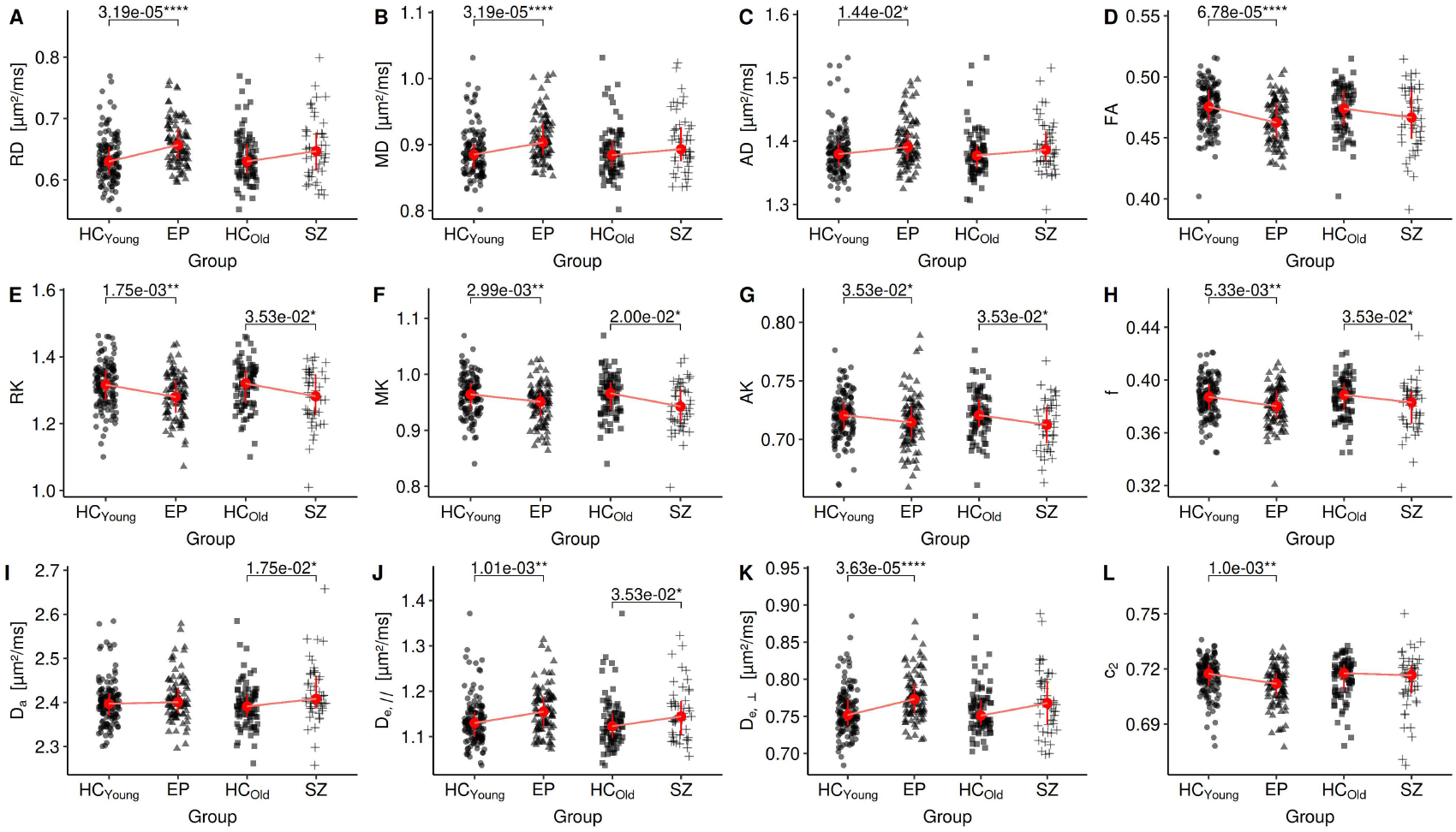
Strip plot of the group comparisons. Each clinical group is compared to its respective HC group. *:p≤5e-2, **:p≤1e-2, ***:p≤1e-3, ****:p≤1e-4.

Differences between SZ and HC_Old_ were more limited, with no significant DTI metric. Kurtosis, however, was lower in SZ (**Fig. 1E-G**; RK: p=.035, MK: p=.020, AK: p=.035). The WMTI-W model revealed reduced *f* (**Fig. 1H**; p=.035), increased *D*_a_ (**Fig. 1I**; p=.017), and *D*_e,∥_ (**Fig. 1J**, p=.035). Effect sizes were moderate (mean |*d*|=.38) with the highest being MK (*r*=-.50), followed by *D*_*a*_ (*r*=.50), RK (*r*=-0.47), and *f* (*r*=-.43) (**Table S3**).

### Microstructure imaging estimates of the white matter JHU ROIs

Group comparison of the JHU ROIs revealed distinct patterns in the microstructure metrics tendencies in both EP and SZ (**Fig. 2**). In EP, significant alterations in microstructure metrics were more widespread than in SZ.

**Figure 2.**
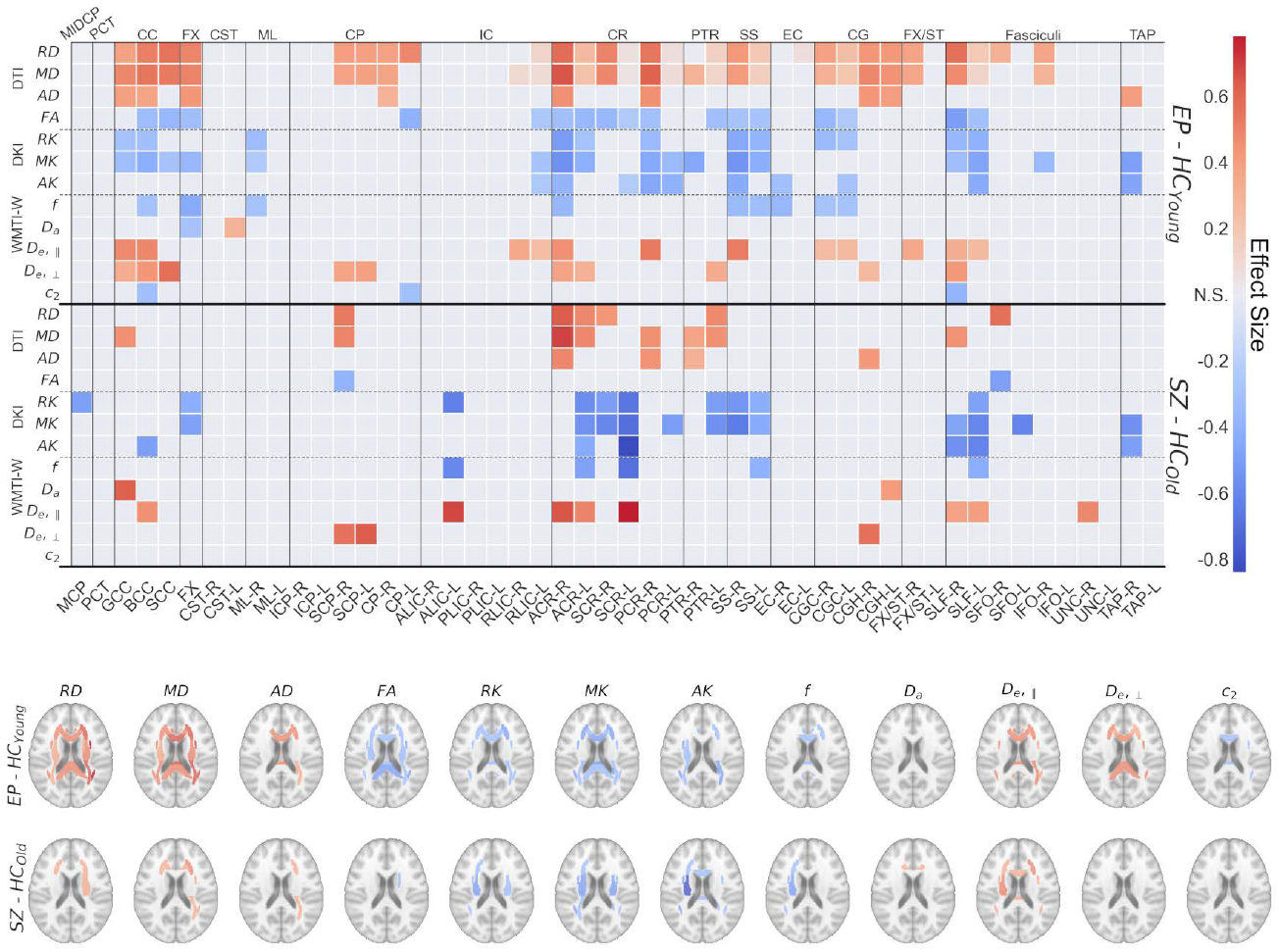
Heatmaps and brainplots of the JHU ROI group comparison. Top: group comparisons (y-axis right) heatmap of the effect size, thresholded for significance, for each dMRI metric (y-axis left) and each region of interest (x-axis). Bottom: brainplots of the same comparisons. The colorbar is shared. Red: clinical group > HC, Blue: clinical group < HC. NS: p>0.05, *:p≤5e-2, **:p≤1e-2, ***:p≤1e-3, ****:p≤1e-4. Numeric p-values and effect sizes can be found in Table S5, S6. *Abbreviations*: L: left; R: right; MCP: Middle cerebellar peduncle; PCT: Pontine crossing tract; GCC/BCC/SCC: Genu/Body/Splenium of corpus callosum; ; FX: Fornix (column and body); CST: Corticospinal tract; ML: Medial lemniscus; ICP/SCP: Inferior/Superior cerebellar peduncle; CP: Cerebral peduncle; ALIC/PLIC/RPIC: Anterior limb/Posterior limb/ Retrolenticular part of internal capsule; ACR/SCR/PCR: Anterior/Superior/Posterior corona radiata; PTR: Posterior thalamic radiation; SS: Sagittal stratum; EC: External capsule; CGC: Cingulum (cingulate gyrus); CGH: Cingulum (hippocampus); FX/ST: Fornix / Stria terminalis; SLF: Superior longitudinal fasciculus; SFO/IFO: Superior/Inferior fronto-occipital fasciculus; UNC: Uncinate fasciculus; TAP: Tapetum.

DTI: In EP, MD and RD showed significant increases in 27 and 28 of the 50 ROIs, followed by lower FA in 17 ROIs. The most affected ROIs were the ACR-R (*d*_*MD*_=.65, *d*_*RD*_=.58), posterior CR-R (PCR, *d*_*MD*_=.61) and SLF-R (*d*_*RD*_=.57, *d*_*FA*_=-.50). In SZ, significant alterations were sparse, only found in 6, 8 and 2 ROIs for MD, RD and FA respectively. The most affected areas were the ACR-R (*d*_*MD*_=.69, *d*_*RD*_=.63), SFO-R (*d*_*RD*_=.57, *d*_*FA*_=-.49) and right cingulum (CGH-R, *d*_*MD*_=.51).

DKI: Kurtosis metrics were lower in both patient groups. In EP, 17 ROIs have significantly lower MK, 12 had lower RK and 10 has lower AK. The most affected ROIs were the ACR-R (*d*_*MK*_=-.56, *d*_*RK*_=-.51), the right sagittal stratum (SS-R, *d*_*MK*_=-.54, *d*_*RK*_=-.45), and right tapetum (TAP-R, *d*_*MK*_=-.49). In SZ, MK was significantly different in 12 ROIs, RK in 10 and AK in 6. The left superior CR had the highest effect size (SCR-L, *d*_*AK*_=-.84, *d*_*MK*_=-.70, and *d*_*RK*_=-.67), followed by SS-R (*d*_*MK*_=-.65) and the left anterior limb of the internal capsule (ALIC-L, *d*_*RK*_=-.63).

WMTI-W: The biophysical model attributed the changes observed in DTI and DKI metrics in EP and SZ to a reduced axonal water fraction *f* (in 9 and 5 ROIs, respectively) but especially higher extra-axonal diffusivities: *D*_*e,∥*_ (in 12 and 8 ROIs) and *D*_*e*,⊥_ (in 10 and 3 ROIs). In EP, the strongest effect sizes for *f, D*_*e,∥*_ and *D*_*e*,⊥_ were achieved respectively in the fornix (FX, *d*_*f*_=-.44), PCR-R (*d*_*De,∥*_=.53), and the splenium of the CC (SCC, *d*_*De*,⊥_=.57). For the same metrics, the SCR-L (*d*_*f*_=-.69, *d*_*De,∥*_=.78), ALIC-L (*d*_*De,∥*_=.68), the left superior cerebellar peduncle (SCP-L, *d*_*De*,⊥_=.63) were the ROIs with highest effect size in SZ.

EP vs SZ: Overall, the Dice coefficient was .36, so 18 significant ROI alterations were common to both group comparisons and we found no differences between groups. The correlation between all ROI effect sizes of EP and SZ (**Fig. S2**) showed strong agreement in MK and MD (*r*>0.6) followed by RK and RD (*r*=.55, .47) and FA (*r*=.41). Furthermore, *D*_*e*,⊥_ had stronger agreement (*r*=.61) than axonal water fraction *f* (*r*=.41), suggesting common features between EP and SZ as altered myelination and thus changes in the extra-axonal environment.

After covarying for the core WM, we found an increased MK, RK and decreased RD, MD in the posterior limb of the internal capsule (PLIC, *d*∼.40) of EP and an increase of the ACR-R MD in SZ.

All the ordered ROI effect sizes for DTI, DKI and WMTI-W can be found in **Table S4**, while a 3D render of the alteration frequency can be found in **Fig. S4, 5**.

### Regional agreement between effect sizes of ENIGMA and Lausanne Psychosis cohort

We evaluated the regional effect sizes found in DTI and DKI metrics for the two groups of our LSP cohort against the regional effect sizes in the review by Kelly et al.^8^ where only effect sizes for FA, MD and RD were reported (**Fig. 3**). It is noteworthy that the ENIGMA meta-analysis reports mainly on SZ. Overall, the agreement depended on the metric and group in question. Regional FA effect size correlations between ENIGMA and LSP were stronger in SZ (*r*=.66, **Fig. 3F**) than EP (*r*=.47, **Fig. 3A**). The regional agreement of MD (**Fig. 3B, H**) and RD (**Fig. 3C, I**) was low-moderate (0.26<*r*<0.41). Instead, MK and RK in the LSP cohort showed the strongest associations (.53<*r*<.67) when compared to ENIGMA MD (**Fig. 3D, J**) and RD effect sizes (**Fig. 3E, K**), respectively.

**Figure 3.**
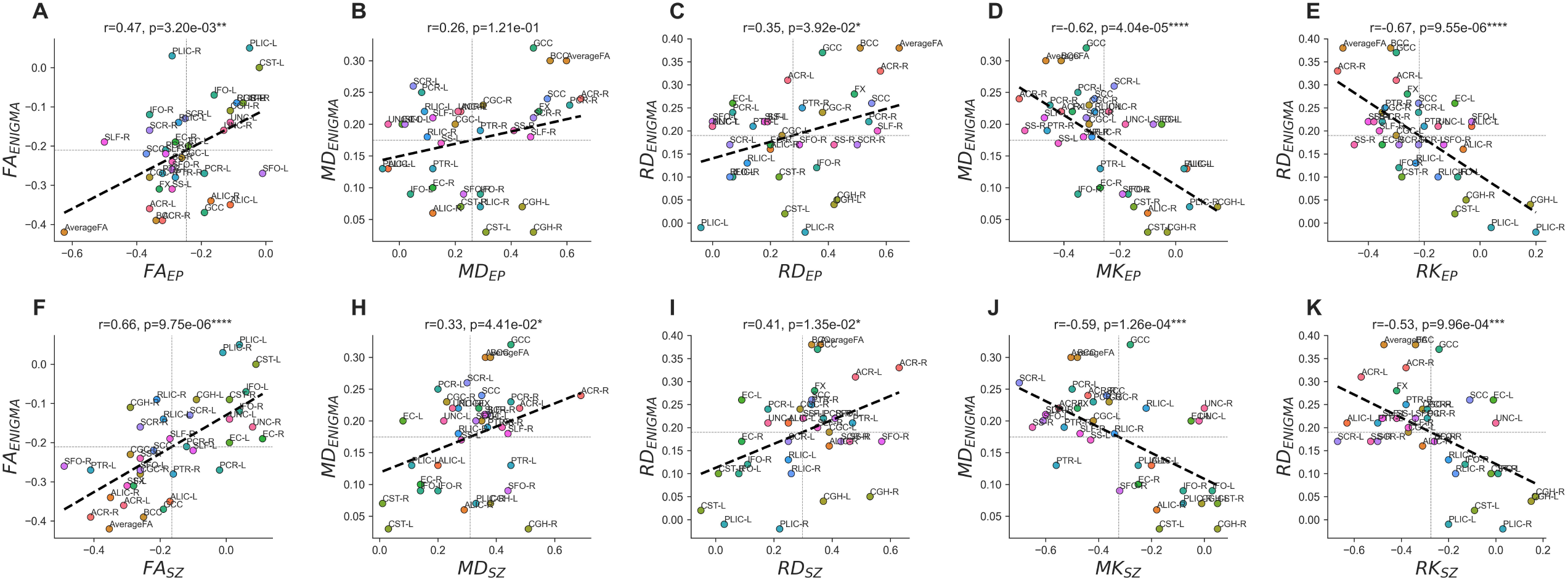
ENIGMA versus Lausanne psychosis (LSP) regional effect sizes (*Cohen’s d*). Plots are divided by EP (top row) and SZ (bottom row). Here, “averageWM” refers to ENIGMA average WM skeleton while for LSP it corresponds to the WM core (average of all JHU ROIs combined in one mask).

### Signal representations versus biophysical model: sensitivity versus specificity

Signal representations (DTI, DKI) yielded a higher number of significant differences between groups than WMTI-W metrics. When averaging regional effect sizes in significant ROIs, in EP (**Fig. 4A**), MK and AD had the strongest effect size (|*d*_*ep*_|=.40) followed by *D*_*e*,⊥_ (*d*_*ep*_=.38). In SZ (**Fig. 4B**), the highest effect sizes were achieved by metrics beyond DTI: *D*_*e*,⊥_ (*d*_*sz*_=.59), AK (*d*_*sz*_=-.57) and MK (*d*_*sz*_=-.56). However, the larger number of significant ROIs in DTI may bias this result. Remarkably though, even when averaging regional effect sizes irrespective of their significance (**Fig. S3**), MK was the metric achieving the highest absolute effect size in SZ (*d*_*sz*_=-.28), over RD (*d*_*sz*_=.26) and MD (*d*_*sz*_=.27). Finally, while signal representations (DTI/DKI) are more sensitive to WM changes than biophysical model parameters (WMTI-W), it is noteworthy that the effect sizes between the two families of features are not dramatically different, while biophysical models retain superior specificity in characterizing pathology.

**Figure 4.**
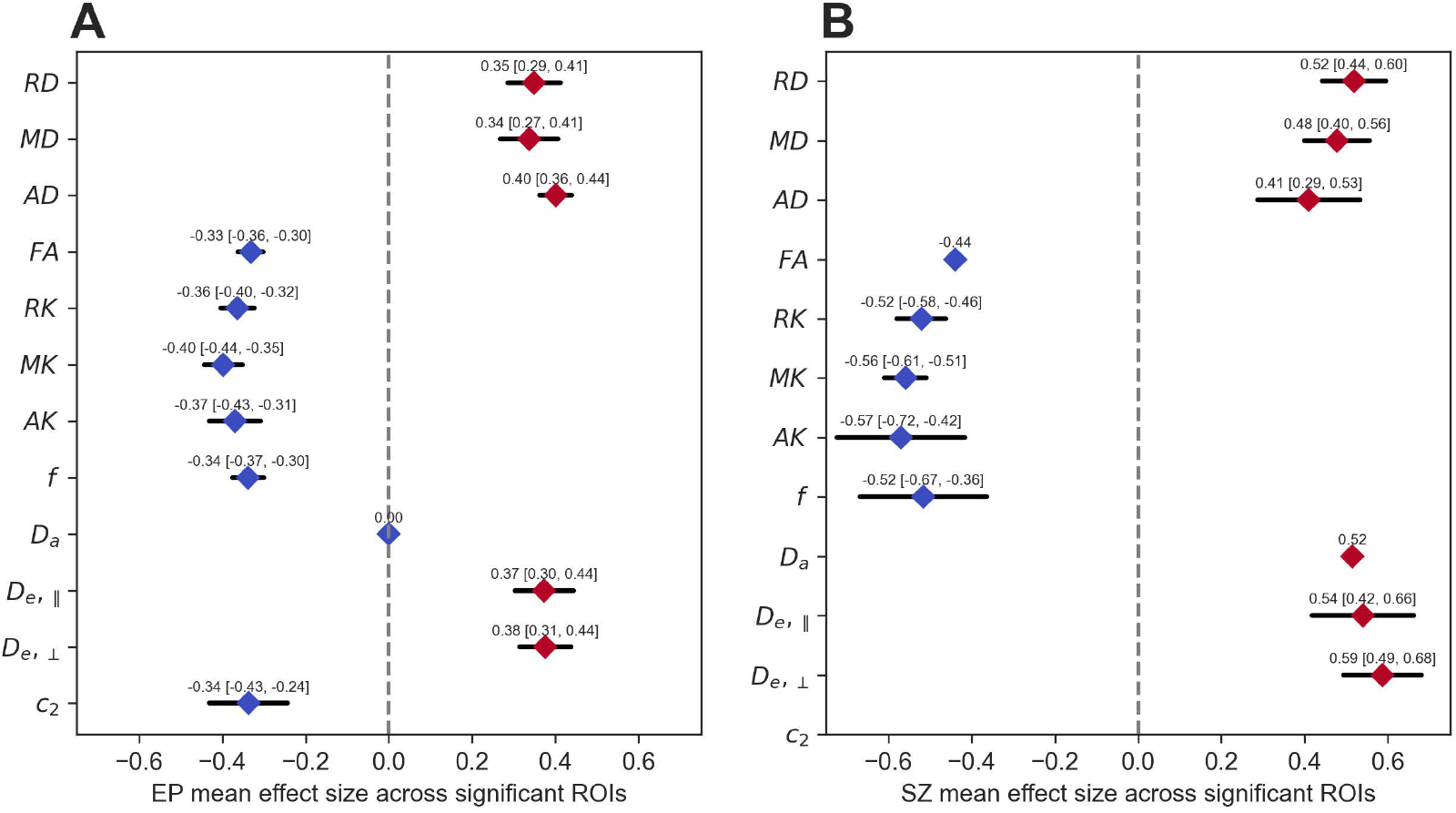
Average and confidence interval of the effect sizes of significant ROIs for each dMRI metric in EP vs HC_Young_ and SZ vs HC_Old_.

### Association between microstructure estimates, clinical characteristics and psychopathological symptom domains

We correlated clinical characteristics to the microstructure in the WM core (i.e., averaged measure across all WM ROIs) because the diffusion MRI metrics showed widespread consistent changes across JHU ROIs in patients and strong effect sizes. Furthermore, this approach limits considerably the number of statistical tests. There were no significant associations between WM microstructure and age at psychosis onset, nor with illness duration (covaried with age). The intra-axonal diffusivity, *D*_*a*_, showed significant negative association with CPZ-equivalent dose in the SZ group (p_FDR_=.013), significantly different from EP slope (p_FDR_=.0019). To better understand the aging trajectories of WM microstructure in patients vs. HC we examined the interaction between age and the two grouping variables (EP+SZ) and HC, but not significant differences were found after FDR correction (**Fig. S6**). The associations between microstructure metrics in the WM core and symptom scores were also examined. Overall, correlations between psychiatric scales and WM microstructure metrics were stronger in SZ than EP and showed different cluster patterns between groups. However, no correlation survived FDR correction (**Fig. S7**).

## Discussion

With the present analysis we studied a rich early psychosis and schizophrenia cohort (LSP), employing more advanced diffusion metrics than DTI, namely DKI and a comprehensive microstructure model, WMTI-W, both aimed at characterizing the WM pathology with increased sensitivity and specificity. We compared the patterns and effect sizes obtained from EP and SZ with the current literature as well as between the two patient groups. Finally, we tested the dMRI metrics against commonly used clinical measures and characteristics.

Our findings revealed that WM alterations are already present and widespread at the EP stage. We found a vast increase in diffusivities in both groups, especially in RD, and decrease in FA and kurtosis in the WM core with large effect sizes. The increase in DTI diffusivities and decrease in FA confirms previous findings in similar cohorts^4,8,23,71^ and high effect sizes in the average WM^8^. The large effect sizes found for DKI metrics, particularly between SZ and HC_Old_, support the added value of going beyond DTI in quantitative diffusion MRI analyses of clinical populations, as supported by several other patients studies^45,50,72^ and animal models of disease^46,47^.

Regionally, the largest effect sizes were observed consistently in the ACR-R, but also in the PCR, the SLF and the CC in EP. The corona radiata has been often reported to be affected in SZ, showing among the strongest regional effect sizes^8^. The CR collects fibers condensing from the cortex to the internal capsule, brainstem and thalamus^73^ and is important for emotion, motivation, cognition processing, and decision-making^74^. Specifically in SZ, reduced FA in the ACR was associated to impaired social cognition^75^, and higher severity of auditory and verbal hallucinations^76^. The SLF is an association fiber that connects the frontal lobe to the rest of the ipsilateral brain, and whose FA increases significantly during normal development^77^. Reduced integrity of the SLF has previously been found in clinical high-risk subjects^78^, recent-onset SZ^79^, and was associated to reduced verbal fluency^79^. A recent meta-analysis of early-onset psychosis^6^ (onset<18y.o.) and ultra high-risk individuals^80^ reported both the SLF and PCR as the main affected ROIs, which contrasted findings in adults^8^. All the high effect size ROIs identified in early-onset and adult patients^6,8^ (SLF, PCR, ACR, CC) also showed the highest effect sizes in our LSP cohort.

We expected a good regional agreement between EP and SZ, which was confirmed by a high Dice score (*dice*=.36) and strong correlation between EP and SZ effect sizes. The regional agreement in effect sizes was particularly high for MD, MK (*r*>.60) and for *D*_*e*,⊥_ (*r*=.61), suggesting higher sensitivity of these metrics at detecting shared regional alterations between early and late stages compared to the other metrics. These results indeed support hypothesis that FA differences to HC originate during the development and adolescence and remain throughout the lifespan^23–26^. In this context, the widespread alterations found in EP could result from a shift in peaks in the WM development, achieved earlier in patients^4,25^ than HC and resulting in a persistent offset of the WM. However, it remains unclear from our data whether the WM alterations worsen with disease progression towards chronic SZ, as effect sizes do become more pronounced in SZ but fewer ROIs are significantly different between SZ and HC_Old_. Other studies, on the other hand, report worsening of the FA trajectories with age at later stages of the disease^4,25,81^. Our SZ cohort size is perhaps somewhat limited with respect to the wide age range, which could lower our statistics for this group.

Regarding agreement of the alterations found in the LSP cohort with the literature^8^, we found an excellent agreement between FA effect sizes in ENIGMA vs both our EP and SZ cohorts (*r*_*EP*_=.47, *r*_*SZ*_=.66), indicating the patterns reported in ENIGMA are reproducible in different cohorts evaluated with different methodologies. Surprisingly, we found better agreement of ENIGMA MD, RD effect sizes with our MK and RK effect sizes (*r*>.60) rather than MD or RD (.26<*r*<.41). A possible explanation is that, in the simple DTI fit that does not estimate kurtosis jointly, the DTI metrics are biased by higher order features of the diffusion signal, i.e. kurtosis arising from the non-gaussian behavior of the water molecules. A full DKI fit improves the accuracy of DTI metrics while properly separating the non-gaussian effects in the kurtosis metrics (MK, RK, AK)^82^. In other words, ENIGMA DTI measures are influenced by a combination of gaussian and non-gaussian diffusion, while the LSP DTI measures are not, which could explain the better correlation between ENIGMA first-order measures (MD, RD) and LSP higher order measures (MK, RK).

When covarying the ROI diffusion measures with the matching core WM measures, only two ROIs remained significant: the PLIC in EP (increased MK, decreased MD) and ACR-R in SZ (increased MD). This result is consistent with ENIGMA^6,8^ previous findings in which only the PLIC (increased FA) survived the same type of analysis. This further confirms that despite the regional specificity, the alterations are driven by a common general effect across WM.

Finally, the lack of significant associations of WM microstructure metrics with age at first psychosis, medication, duration of illness or symptomatology is also aligned with the literature^8,83,84^. Specifically, regarding symptomatology, the lack of associations could be explained by its transitory nature. The patient’s symptomatologic state at the time of the MRI scan may differ from the state that led to the assigned clinical score. Furthermore, the difference in cluster patterns between the EP and SZ stages may be due to sampling variability and heterogeneity between the two cohorts rather than specific symptoms-microstructure relationships.

For the first time, we used WMTI-W, a comprehensive biophysical model of WM, to tease apart possible pathological contributions to the reported WM differences. Here, the effect sizes carried by the WMTI-W parameters were marginally lower than those by DTI or DKI. Nonetheless, the changes in model metrics improve the interpretation of WM changes in EP and SZ. Biophysical models come with improved specificity at a moderate cost in sensitivity^85^. For example, using a similar WM model in stroke showed preferential changes as increased *f* and dramatically decreased *D*_*a*_, consistent with cytotoxic edema and beading^72^, in multiple sclerosis as decrease in *f*, fiber alignment and higher extra-axonal diffusivities^72^, consistent with demyelination, and in a rat model of Alzheimer’s disease as a decrease in *f* and *D*_*a*_, consistent with axonal injury and loss^49^.

In EP and SZ, WMTI-W helps attribute the observed alterations to the extra-axonal compartment, due to the significant increase in *D*_*e,∥*_ and *D*_*e*,⊥_, and the lower axonal water fraction *f*. In validation studies, a similar – albeit more pronounced – pattern of alteration (*f* ↓, *D*_*e,∥*_ ↑, *D*_*e*,⊥_ ↑) has been shown to arise from demyelination, induced via chronic cuprizone intoxication in mice^46,47,86^ (**Table 1**). Ultrastructural post-mortem studies in SZ reported myelinated WM fiber pathology in form of decompacting and splitting of the myelin sheath and inclusions of vacuoles in-between myelin layers, small-axons atrophy, and the presence of swollen or dystrophic oligodendroglia^11–13^, all of which can explain the demyelination-like pattern drawn by *D*_*e,∥*_, *D*_*e*,⊥_ and *f*. In the SZ group the WM core analysis also suggested higher intra-axonal diffusivity, *D*_*a*_, in this group, but only two specific ROIs sustained this trend (CING and CC).

Previous works using NODDI^34^, a comparable but more constrained biophysical model, also reported reduced neurite density (comparable to *f*) in several ROIs and increased orientation dispersion index (corresponding to lower *c*_2_) in both first episode and chronic SZ^87^. These patterns of preferentially altered extra-axonal environment are also consistent with reports of global increase in “free water” using the FWI technique in SZ cohorts^35^, although FWI conflates potential pathological mechanisms by defining a tissue compartment (intra- and extra-axonal) vs a free water (CSF) one.

### Limitations

The first two main limitations of the present study are the use of the age-range approach instead of age-matching and the use of data from two scanners that is a source of bias, possibly even after careful harmonization. The age-range approach was preferred to maintain the sample size as large as possible to better correct for the scanner effect and increase the statistical power of the analyses. In addition, we believe it was preferable not to pair participants by age at the cost of pairing data from different scanners, which could introduce more bias. With regards to the WM biophysical model, the modeling of the axonal dispersion as Watson distribution is not ideally suited for multi-fiber configurations, e.g. crossing fibers^16^. Finally, while the subject numbers in the cohort are sufficiently high to support our conclusions, they are not balanced between groups, which is due to the concrete challenge in recruiting EP and SZ patients. In the light of these limitations, the generalizability of our conclusions needs to be confirmed in other cohorts.

### Conclusions

In conclusion, with this work we confirmed that WM alterations are already present at the early psychosis stages, consistent with shifted maturation, and are more widespread in EP but more pronounced in SZ. The regional alterations of higher DTI diffusivities and lower kurtosis found in our cohort matched the DTI patterns found in ENIGMA-schizophrenia. WMTI-W model parameters indicate that the WM alterations in patients manifest preferentially perpendicular to the axons as extra-cellular increase in diffusivities, decreased intra-cellular water fraction, and loss of fiber alignment, consistent with early myelin aberrations. The direct link between WM alterations and symptomatology is, however, limited.

## Data Availability

Data is not available for public use.

## Acknowledgments

This work was supported by the Swiss National Science Foundation (PCEFP2_194260, to I.J.), the National Center of Competence in Research (NCCR) “SYNAPSY - The Synaptic Bases of Mental Diseases” from the Swiss National Science Foundation (n° 51NF40 – 185897 to KQD & PC) and the Foundation Alamaya. LA is supported by Carigest fellowship. PK, DD and LA are supported by the Adrian & Simone Frutiger Foundation.

