## Supplementary Material for "White Matter Microstructure Alterations in Early Psychosis and Schizophrenia"

#### Methods details

##### Acquisition details

Magnetization-prepared rapid acquisition gradient echo (MPRAGE): echo time (TE) 2.98ms, repetition time (TR) 2300 ms, inversion time (TI) 900 ms, field of view (FOV) 160 x 240 x 256 $mm^3$ and voxel size 1 x 1 x 1.2 $mm^3$, acquisition time 7 minutes. Pulsed Gradient Spin-echo (PGSE) echo planar imaging (EPI) sequence: acquired in cartesian q-space coverage totalling 129 (Prisma) or 257 (Trio) DWI volumes. Prisma: TE = 144 ms, TR = 6.1 s; Trio: TE = 103 ms, TR = 5.9 s. FOV = 211 x 211 x 114$mm^3$, voxel size = 2.2 x 2.2 x 3$mm^3$, 96x96x38 slices, partial Fourier = 0.75, 1 b0 acquisition 128 (Prisma) and 256 (Trio) directions, acquisition time 13 minutes. Any participants with pacemaker, cochlear implant, implant of stimulator or drug pump, glucose sensor, bypass valve or pregnant were not allowed in the MRI scanner and excluded from the study.

##### Preprocessing details

The diffusion preprocessing pipeline included MP-PCA denoising (Veraart et al., 2016) and Gibbs ringing correction (Kellner et al., 2016; Lee et al., 2021). The EPI distortions were corrected using ANTs non-linear registration since no reverse phase encode image or magnetic field map were acquired. The registration of the DSI b=0 volume to the MPRAGE was constrained only in the direction of the distortion, for then warping the estimated correction back to each DWI volume in native space (Alemán-Gómez et al., 2023; Tax et al., 2022). Thereafter, the distortion-corrected images were further corrected for eddy currents and motion using FSL *eddy* (Andersson and Sotiropoulos, 2016). Since FSL eddy does not support DSI data natively (see *–data_is_shelled* in: https://fsl.fmrib.ox.ac.uk/fsl/fslwiki/eddy/UsersGuide), temporarily merging or directly dropping a subset of volumes based on their b-values in order to simulate shells was necessary to accommodate the algorithm. Namely, volumes b=1500 and b=2000 were merged as b=1750, b=4000 was merged to b=4500 while b=6000 and b=8000 were removed. Right after the eddy execution the merged volumes were split back into their original b-values except for the dropped volumes (for the dMRI metric estimations only the b-values≤2500 were needed). The reduced DWI images featured 29 and 57 directions (Prisma, 1:b=0, 3:b=500, 6:b=1000, 4:b=1500, 3:b=2000, 12:b=2500; Trio, 1:b=0, 6:b=500, 12:b=1000, 8:b1500, 6:b=2000, 24:b=2500). Data quality was assessed by visual inspection and automated means (FSL's eddy QUAD and SQUAD (Bastiani et al., 2019)). The estimated quality metrics were volume-to-volume (absolute movement) and within-volume motion (relative movement), eddy current-induced distortions and data outliers. For the Trio scanner the study mean absolute movement was $1.08 \pm 0.39$ mm (min:0.21, max:3.21); while the mean relative movement was $0.02 \pm 0.013$ mm (min:0, max:0.1). For the Prisma scanner the study mean absolute movement was $0.46 \pm 0.31$ mm (min:0.13, max:3.36); while the mean relative movement was $0.1 \pm 0.06$ mm (min:0.02, max:0.61).

**Age correction**

Significant differences in age were found between patients and control groups. To better model age and correct for its effect, five different model were evaluated for each of the 12 WM core metric: no age effect (intercept-only), linear age effect (LAE), quadratic age effect (QAE), linear age effects with group interaction (LAEG) and quadratic age effects with group interaction (QAEG); for a total of 60 models. The model selection criteria was to minimize the Bayesian information criterion (BIC) between the fitted model for one metric, and then selecting the most popular correction model among the metrics. The procedure returned the QAE as the best model (7 metrics), followed by LAE (3; FA, $D_a$, $D_{e,\perp}$), and intercept-only (2). The intercept-only model resulted from the RK and $f$ models. In $f$ the second best model was the QAE (intercept-only BIC: -1861 < QAE: -1860 < LAE: -1858), while, in RK, LAE and QAE were very close (intercept-only: -813.14 ≤ LAE: -809.13 ∼ QAE: -809.04). Thus, we concluded that the QAE was the best model to correct for age effects in our cohort. See Figure S1 for an example.

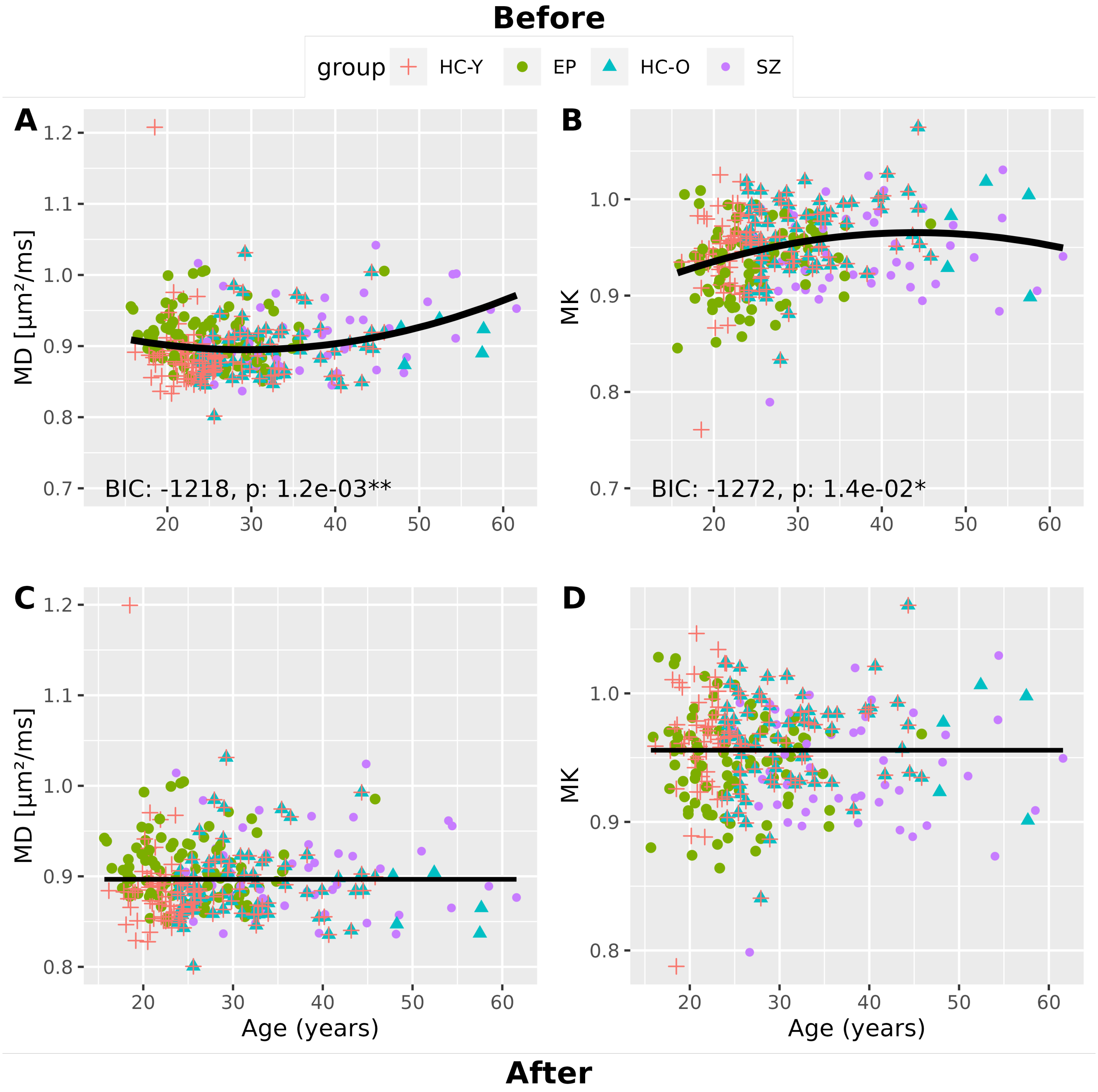


Figure S 1: Before (A, B) vs after (C, D) examples of fits for the best age correction model of the WM cores MD (A, C) and MK (B, D). The procedure selected the quadratic age effect (QAE) as best model. In the before-plots (A, B), the black solid line indicates the predicted values of MD or MK given the corresponding age. In the after age correction plots (C, D), the model intercept is maintained and just the age effect is removed.

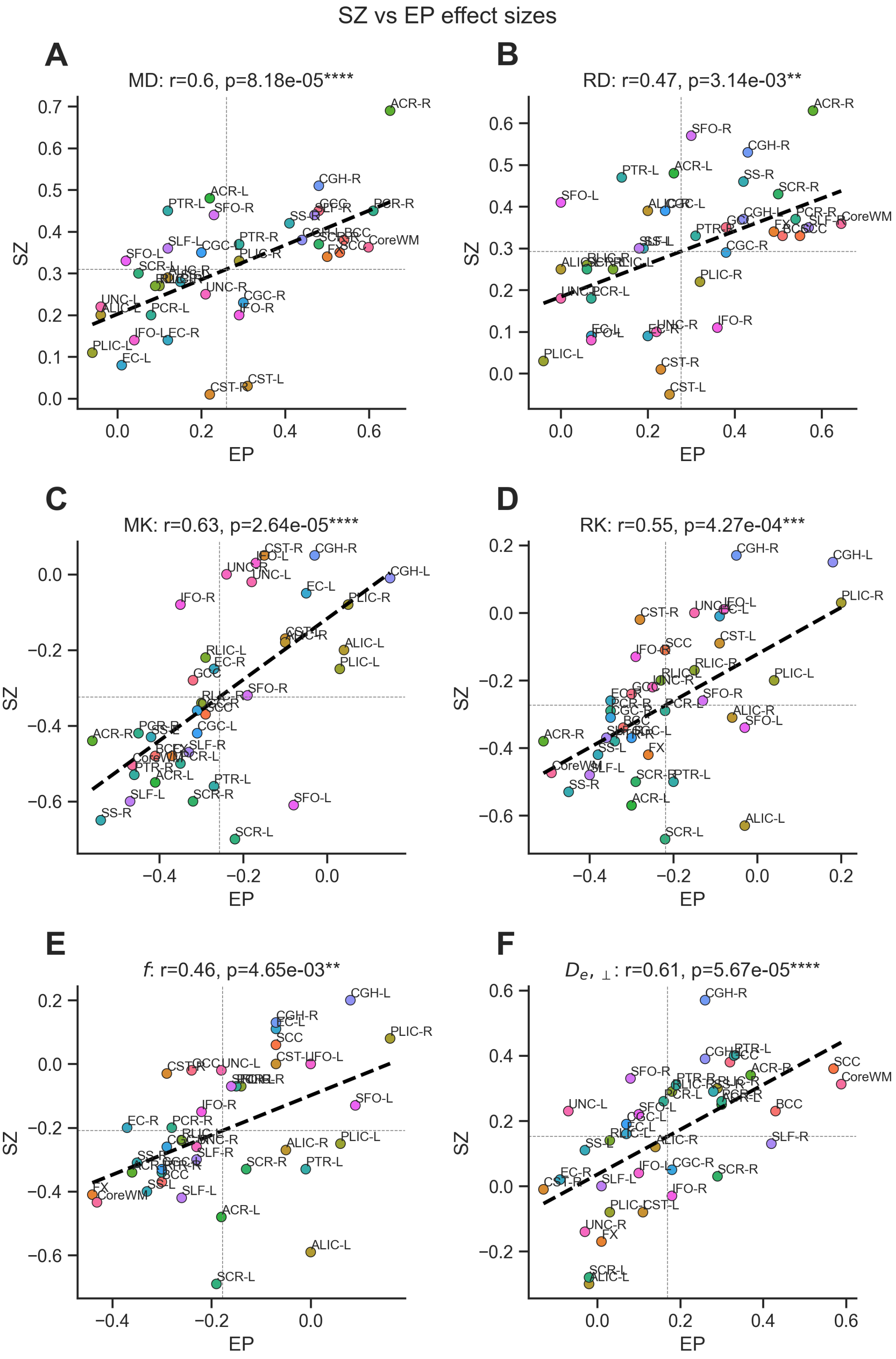


Figure S 2: Scatterplot of regional effect sizes of EP vs SZ. A moderate-to-strong correlations were found between EP and SZ in MD, RD, MK, RK, $f$ and $D_{e,\perp}$ (0.47<r<0.63). The correlation showed stronger agreement (r>0.6, A and C) in MK and MD than RK and RD (r=.55, .47, B and D) and FA (r=.41). Furthermore, the agreement in $D_{e,\perp}$ attributes stronger common features between EP and SZ in the extra axonal environment (r=.61, F) rather than in the axonal fraction, f (r=.41, E)

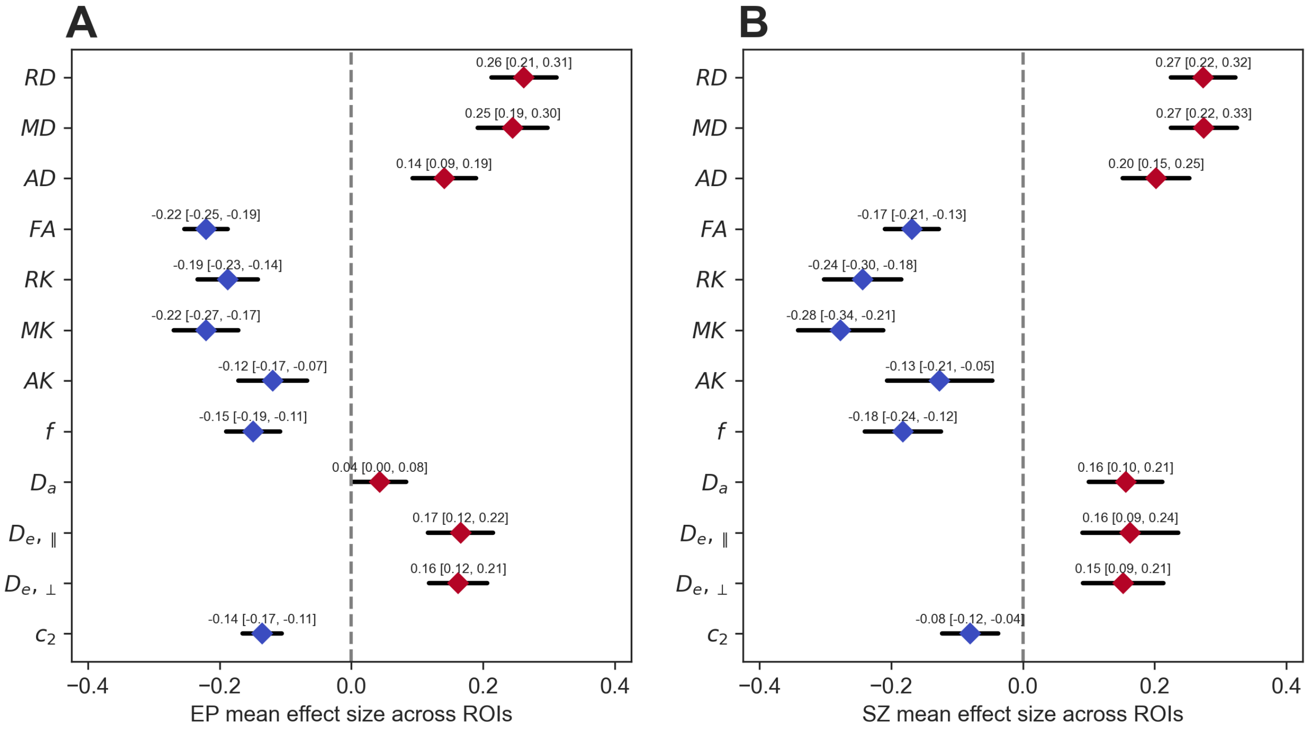


Figure S 3: Forest-like plot of average effect size across ROI per clinical group.

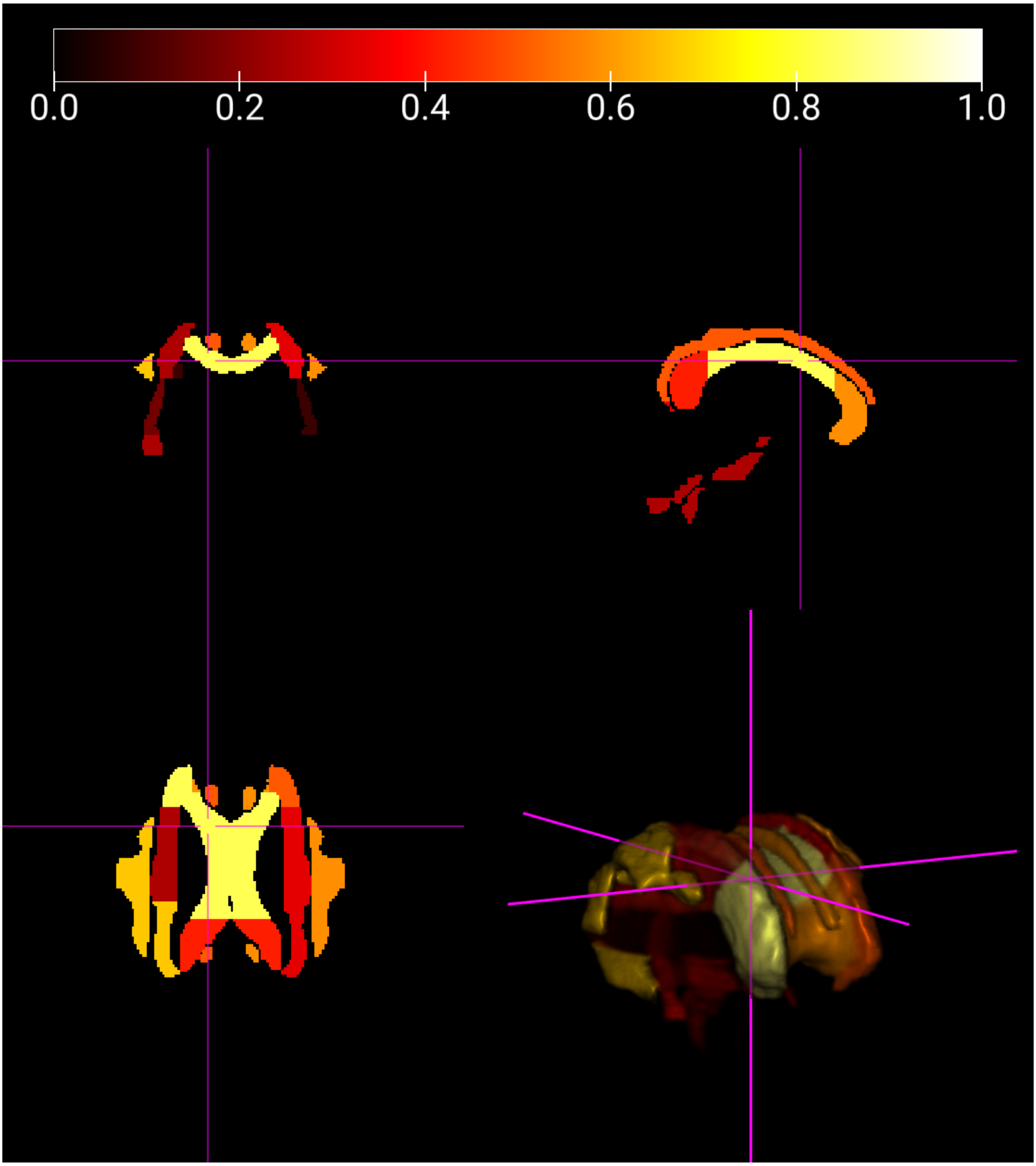


Figure S 4: Render of the JHU atlas alterations for the EP group. Color code represent the frequency of alterations across the 12 dMRI metrics (0=no alterations, 1= all metrics detected alterations).

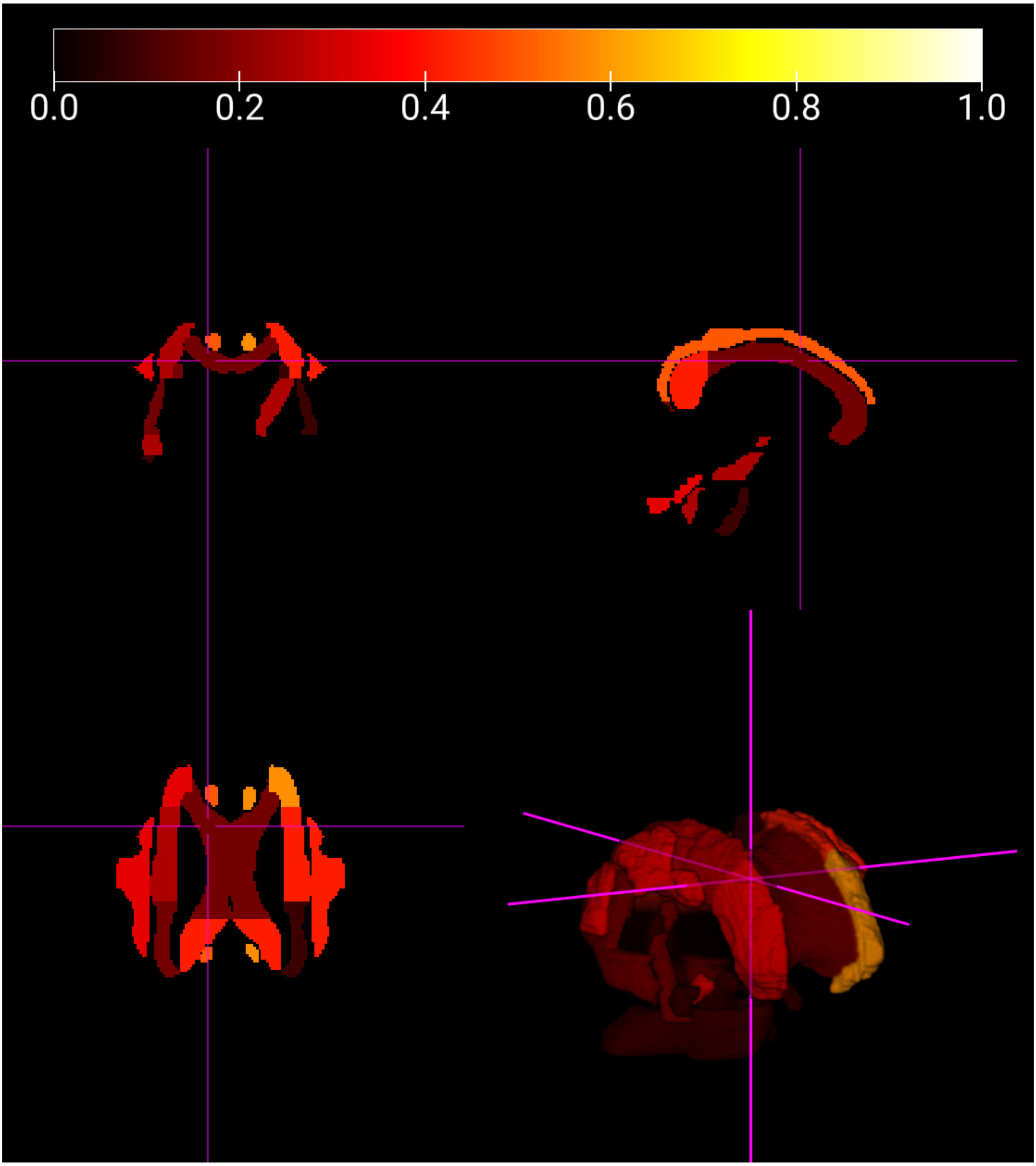


Figure S 5: Render of the JHU atlas alterations for the EP group. Color code represent the frequency of alterations across the 12 dMRI metrics (0=no alterations, 1= all metrics detected alterations).

### Microstructure estimates and Clinical covariate analysis

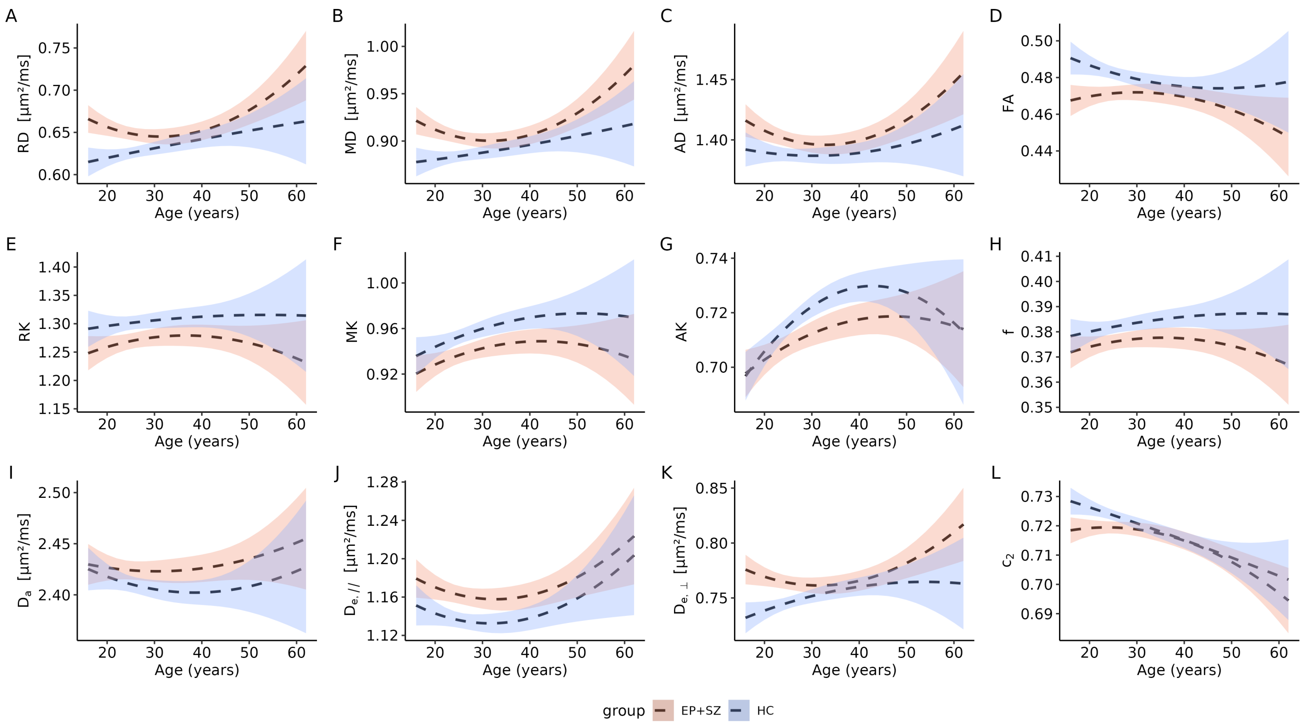


Figure S 6: Aging trajectories of the estimated white matter cores microstructures metrics. The dashed black lines with colored confidence intervals are the quadratic regressions that encompass all the patients (EP+SZ, light orange confidence interval) and all healthy controls (HC, light blue confidence interval) together. Contrasts between HC and PT of RD, MD, FA and $D_{e,||}$ were significant before FDR correction.

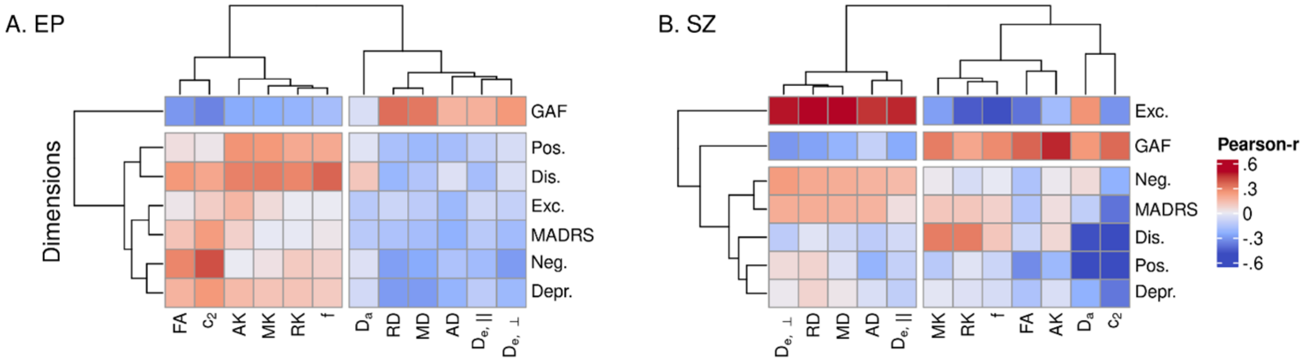


Figure S 7: Hierarchical clustering of the correlation matrices between WM microstructure estimates and psychopathological symptoms domains. No significant findings survived FDR correction. A: EP; B: SZ. Microstructure estimates were corrected for sex and quadratic age. The line diagrams indicate the clustering results. GAF: Global Assessment of Functioning scale; MADRS: Montgomery-Asberg Depression Rating Scale; Positive and Negative Syndrome Scales: pos.: positive; neg.: negative, dis.: disorganized; depr.: depressive; exc.: excited.

#### Diagnosis

| | | EP (N=93) | | SZ (N=47) | |
|---|---|---|---|---|---|
| | | N | Pct. | N | Pct. |
| Diagnosis | Bipolar disorder | 3 | 3.2 | 0 | 0.0 |
| | Bipolar disorder with psychotic features | 4 | 4.3 | 0 | 0.0 |
| | Brief psychotic disorder | 10 | 10.8 | 0 | 0.0 |
| | Cannabis-induced psychotic disorder | 1 | 1.1 | 0 | 0.0 |
| | Delusional disorder | 0 | 0.0 | 1 | 2.1 |
| | Depressive disorder with psychotic features | 2 | 2.2 | 0 | 0.0 |
| | Schizoaffective disorder | 9 | 9.7 | 4 | 8.5 |
| | Schizophrenia | 53 | 57.0 | 42 | 89.4 |
| | Schizophreniform disorder | 9 | 9.7 | 0 | 0.0 |
| | Schizotypal personality disorder | 1 | 1.1 | 0 | 0.0 |
| | Unspecified psychotic disorder; Depressive disorder | 1 | 1.1 | 0 | 0.0 |

Table S 1: Early psychosis and schizophrenia patients individual diagnosis. EP early psychosis, SZ chronic schizophrenia, Pct. Percentage

### White matter core comparisons

| metric | $p_{FDR}$ | | effsize | CI low | CI high |
|---|---|---|---|---|---|
| $RD$ [µm²/ms] | $3.2 \times 10^{-5}$ | **** | 0.64 | 0.37 | 0.92 |
| $FA$ | $6.8 \times 10^{-5}$ | **** | −0.62 | −0.90 | −0.35 |
| $MD$ [µm²/ms] | $3.2 \times 10^{-5}$ | **** | 0.60 | 0.33 | 0.87 |
| $D_{e,\perp}$ [µm²/ms] | $3.6 \times 10^{-5}$ | **** | 0.59 | 0.32 | 0.86 |
| $RK$ | $1.7 \times 10^{-3}$ | ** | −0.49 | −0.76 | −0.22 |
| $c_2$ | $1.0 \times 10^{-3}$ | ** | −0.49 | −0.76 | −0.22 |
| $MK$ | $3.0 \times 10^{-3}$ | ** | −0.46 | −0.73 | −0.19 |
| $D_{e,//}$ [µm²/ms] | $1.0 \times 10^{-3}$ | ** | 0.45 | 0.18 | 0.72 |
| $f$ | $5.3 \times 10^{-3}$ | ** | −0.43 | −0.70 | −0.16 |
| $AD$ [µm²/ms] | $1.4 \times 10^{-2}$ | * | 0.33 | 0.07 | 0.60 |
| $AK$ | $3.5 \times 10^{-2}$ | * | −0.28 | −0.54 | −0.01 |
| $D_a$ [µm²/ms] | $2.1 \times 10^{-1}$ | ns | 0.16 | −0.10 | 0.43 |

Table S 2: EP vs HC, JHU core WM group comparison, sorted by absolute effect size. CI: confidence interval, FDR: false discovery rate.

| metric | $p_{FDR}$ | | effsize | CI low | CI high |
|---|---|---|---|---|---|
| $MK$ | 0.020 | * | −0.50 | −0.87 | −0.14 |
| $D_a$ [µm²/ms] | 0.017 | * | 0.50 | 0.14 | 0.86 |
| $RK$ | 0.035 | * | −0.47 | −0.83 | −0.11 |
| $AK$ | 0.035 | * | −0.43 | −0.79 | −0.07 |
| $f$ | 0.035 | * | −0.43 | −0.79 | −0.07 |
| $D_{e,//}$ [µm²/ms] | 0.035 | * | 0.40 | 0.04 | 0.76 |
| $RD$ [µm²/ms] | 0.087 | ns | 0.36 | 0.00 | 0.72 |
| $MD$ [µm²/ms] | 0.087 | ns | 0.36 | 0.00 | 0.72 |
| $FA$ | 0.107 | ns | −0.35 | −0.71 | 0.01 |
| $D_{e,\perp}$ [µm²/ms] | 0.124 | ns | 0.31 | −0.05 | 0.67 |
| $AD$ [µm²/ms] | 0.124 | ns | 0.29 | −0.06 | 0.65 |
| $c_2$ | 0.55 | ns | −0.16 | −0.52 | 0.20 |

Table S 3: SZ vs HC, JHU core WM group comparison, sorted by absolute effect size. CI: confidence interval, FDR: false discovery rate.

| | $rd-EP$ | | $md-EP$ | | $ad-EP$ | | $fa-EP$ | | $rk-EP$ | | $mk-EP$ | |
|---|---|---|---|---|---|---|---|---|---|---|---|---|
| 1 | PLIC-L | -0.04 | PLIC-L | -0.06 | ALIC-L | -0.10 | SLF-R | -0.50 | ACR-R | -0.51 | ACR-R | -0.56 |
| 2 | ICP-R | 0.00 | ALIC-L | -0.04 | PLIC-L | -0.09 | CP-L | -0.40 | SS-R | -0.45 | SS-R | -0.54 |
| 3 | ALIC-L | -0.00 | UNC-L | -0.04 | UNC-L | -0.08 | SCC | -0.37 | SLF-L | -0.40 | TAP-R | -0.49 |
| 4 | UNC-L | -0.00 | ICP-R | -0.01 | EC-L | -0.07 | SCR-R | -0.36 | SS-L | -0.38 | SLF-L | -0.47 |
| 5 | SFO-L | 0.00 | EC-L | 0.01 | EC-R | -0.03 | CGC-R | -0.36 | SLF-R | -0.36 | PTR-R | -0.46 |
| 6 | PCT | 0.05 | SFO-L | 0.02 | SLF-L | -0.03 | IFO-R | -0.36 | EC-R | -0.35 | PCR-R | -0.45 |
| 7 | SCR-L | 0.06 | IFO-L | 0.04 | ICP-R | -0.03 | ACR-L | -0.36 | CGC-R | -0.35 | SS-L | -0.42 |
| 8 | RLIC-R | 0.06 | PCT | 0.05 | IFO-L | -0.03 | BCC | -0.34 | PCR-R | -0.35 | BCC | -0.41 |
| 9 | IFO-L | 0.07 | SCR-L | 0.05 | ALIC-R | -0.02 | FX | -0.33 | ML-R | -0.34 | ACR-L | -0.41 |
| 10 | EC-L | 0.07 | PCR-L | 0.08 | ICP-L | -0.01 | ACR-R | -0.32 | PTR-R | -0.34 | FX | -0.37 |
| 11 | PCR-L | 0.07 | ICP-L | 0.08 | CGC-L | 0.00 | PTR-L | -0.32 | BCC | -0.32 | PCR-L | -0.35 |
| 12 | FX/ST-L | 0.12 | FX/ST-L | 0.09 | RLIC-L | -0.00 | PCR-R | -0.31 | TAP-R | -0.31 | IFO-R | -0.35 |
| 13 | RLIC-L | 0.12 | RLIC-L | 0.09 | SCR-L | -0.00 | FX/ST-R | -0.31 | ACR-L | -0.30 | SLF-R | -0.33 |
| 14 | ICP-L | 0.12 | RLIC-R | 0.10 | SFO-R | 0.01 | SLF-L | -0.31 | GCC | -0.30 | SCR-R | -0.32 |
| 15 | PTR-L | 0.14 | PTR-L | 0.12 | CGC-R | 0.01 | SS-R | -0.29 | CGC-L | -0.30 | GCC | -0.32 |
| 16 | ML-L | 0.18 | ALIC-R | 0.12 | IFO-R | 0.02 | SS-L | -0.29 | SCR-R | -0.29 | CGC-R | -0.31 |
| 17 | SLF-L | 0.18 | SLF-L | 0.12 | FX/ST-L | 0.02 | SFO-R | -0.29 | IFO-R | -0.29 | CGC-L | -0.31 |
| 18 | SS-L | 0.19 | EC-R | 0.12 | SFO-L | 0.03 | PLIC-R | -0.29 | CST-R | -0.28 | RLIC-R | -0.30 |
| 19 | TAP-L | 0.20 | ML-L | 0.13 | PCT | 0.03 | SCP-L | -0.28 | FX | -0.26 | RLIC-L | -0.29 |
| 20 | EC-R | 0.20 | ML-R | 0.15 | SS-L | 0.04 | EC-R | -0.28 | ML-L | -0.26 | SCC | -0.29 |
| 21 | ML-R | 0.20 | SS-L | 0.15 | CP-L | 0.04 | PTR-R | -0.28 | UNC-R | -0.25 | PTR-L | -0.27 |
| 22 | ALIC-R | 0.20 | CGC-L | 0.20 | PTR-L | 0.05 | RLIC-L | -0.27 | RLIC-L | -0.23 | EC-R | -0.27 |
| 23 | UNC-R | 0.22 | UNC-R | 0.21 | ML-R | 0.06 | SCP-R | -0.27 | SCC | -0.22 | ML-R | -0.25 |
| 24 | CST-R | 0.23 | TAP-L | 0.22 | PCR-L | 0.06 | CGC-L | -0.26 | FX/ST-R | -0.22 | UNC-R | -0.24 |
| 25 | CGC-L | 0.24 | ACR-L | 0.22 | ML-L | 0.07 | SCR-L | -0.25 | SCR-L | -0.22 | TAP-L | -0.24 |
| 26 | CST-L | 0.25 | CST-R | 0.22 | UNC-R | 0.08 | FX/ST-L | -0.24 | PCR-L | -0.22 | SCR-L | -0.22 |
| 27 | ACR-L | 0.26 | SFO-R | 0.23 | ACR-L | 0.10 | EC-L | -0.24 | PTR-L | -0.20 | FX/ST-R | -0.22 |
| 28 | MCP | 0.28 | PTR-R | 0.29 | PLIC-R | 0.12 | GCC | -0.19 | FX/ST-L | -0.19 | ML-L | -0.20 |
| 29 | SFO-R | 0.30 | IFO-R | 0.29 | RLIC-R | 0.13 | PCR-L | -0.19 | CP-R | -0.17 | SCP-L | -0.20 |
| 30 | PTR-R | 0.31 | PLIC-R | 0.29 | SLF-R | 0.14 | CP-R | -0.18 | UNC-L | -0.15 | SFO-R | -0.19 |
| 31 | PLIC-R | 0.32 | CGC-R | 0.30 | CST-R | 0.16 | ALIC-R | -0.17 | RLIC-R | -0.15 | UNC-L | -0.18 |
| 32 | TAP-R | 0.35 | MCP | 0.31 | PTR-R | 0.18 | ML-R | -0.16 | MCP | -0.13 | IFO-L | -0.17 |
| 33 | IFO-R | 0.36 | CST-L | 0.31 | SCR-R | 0.22 | IFO-L | -0.16 | SFO-R | -0.13 | CST-R | -0.15 |
| 34 | FX/ST-R | 0.37 | CP-L | 0.32 | TAP-L | 0.23 | TAP-R | -0.16 | SCP-R | -0.12 | FX/ST-L | -0.15 |
| 35 | CP-R | 0.38 | FX/ST-R | 0.36 | SCP-R | 0.27 | ML-L | -0.15 | SCP-L | -0.09 | MCP | -0.15 |
| 36 | GCC | 0.38 | CP-R | 0.37 | SCC | 0.27 | UNC-R | -0.13 | CST-L | -0.09 | CP-R | -0.11 |
| 37 | CGC-R | 0.38 | TAP-R | 0.38 | CP-R | 0.29 | ICP-L | -0.12 | TAP-L | -0.09 | CST-L | -0.10 |
| 38 | SCP-L | 0.39 | SCP-R | 0.38 | SCP-L | 0.29 | UNC-L | -0.11 | EC-L | -0.09 | ALIC-R | -0.10 |
| 39 | SCP-R | 0.40 | SCP-L | 0.39 | SS-R | 0.30 | CGH-R | -0.11 | CP-L | -0.09 | SFO-L | -0.08 |
| 40 | SS-R | 0.42 | SS-R | 0.41 | FX/ST-R | 0.31 | ALIC-L | -0.11 | IFO-L | -0.08 | SCP-R | -0.07 |
| 41 | CGH-L | 0.42 | CGH-L | 0.44 | CST-L | 0.34 | ICP-R | -0.10 | ALIC-R | -0.06 | ICP-R | -0.05 |
| 42 | CGH-R | 0.43 | SLF-R | 0.47 | MCP | 0.35 | RLIC-R | -0.09 | CGH-R | -0.05 | EC-L | -0.05 |
| 43 | CP-L | 0.48 | SCR-R | 0.48 | BCC | 0.37 | CGH-L | -0.09 | SFO-L | -0.03 | CGH-R | -0.03 |
| 44 | FX | 0.49 | CGH-R | 0.48 | GCC | 0.39 | PCT | -0.07 | ALIC-L | -0.03 | CP-L | -0.01 |
| 45 | SCR-R | 0.50 | GCC | 0.48 | TAP-R | 0.40 | CST-R | -0.07 | ICP-R | -0.01 | ICP-L | 0.00 |
| 46 | BCC | 0.51 | FX | 0.50 | CGH-L | 0.40 | TAP-L | -0.06 | ICP-L | 0.03 | PLIC-L | 0.03 |
| 47 | PCR-R | 0.54 | SCC | 0.53 | FX | 0.43 | MCP | -0.05 | PLIC-L | 0.04 | ALIC-L | 0.04 |
| 48 | SCC | 0.55 | BCC | 0.54 | CGH-R | 0.43 | PLIC-L | -0.05 | CGH-L | 0.18 | PLIC-R | 0.05 |
| 49 | SLF-R | 0.57 | PCR-R | 0.61 | PCR-R | 0.45 | CST-L | -0.02 | PLIC-R | 0.20 | PCT | 0.14 |
| 50 | ACR-R | 0.58 | ACR-R | 0.65 | ACR-R | 0.45 | SFO-L | -0.01 | PCT | 0.22 | CGH-L | 0.15 |

| | $ak-EP$ | | $f-EP$ | | $da-EP$ | | $depar-EP$ | | $deperp-EP$ | | $c2-EP$ | |
|---|---|---|---|---|---|---|---|---|---|---|---|---|
| 1 | TAP-R | -0.47 | FX | -0.44 | FX | -0.29 | TAP-L | -0.23 | CST-R | -0.13 | SLF-R | -0.38 |
| 2 | PCR-R | -0.46 | EC-R | -0.37 | EC-R | -0.21 | FX | -0.20 | EC-R | -0.09 | BCC | -0.32 |
| 3 | SLF-L | -0.43 | ACR-R | -0.36 | ICP-L | -0.19 | IFO-L | -0.08 | UNC-L | -0.07 | CP-L | -0.31 |
| 4 | SS-R | -0.43 | SS-R | -0.35 | PTR-R | -0.17 | ALIC-R | -0.05 | TAP-L | -0.06 | EC-R | -0.28 |
| 5 | PCR-L | -0.40 | SS-L | -0.33 | CP-L | -0.16 | EC-L | -0.04 | UNC-R | -0.03 | PCR-R | -0.27 |
| 6 | ACR-R | -0.39 | PTR-R | -0.30 | FX/ST-R | -0.15 | ICP-R | -0.02 | SS-L | -0.03 | SS-L | -0.27 |
| 7 | EC-R | -0.32 | ML-R | -0.30 | UNC-L | -0.15 | UNC-L | -0.01 | ALIC-L | -0.02 | FX | -0.27 |
| 8 | PTR-R | -0.30 | BCC | -0.30 | UNC-R | -0.11 | PCT | -0.01 | SCR-L | -0.02 | PLIC-R | -0.27 |
| 9 | CGC-L | -0.30 | CGC-L | -0.30 | EC-L | -0.09 | ALIC-L | 0.00 | FX | 0.01 | ACR-L | -0.25 |
| 10 | FX | -0.28 | CGC-R | -0.29 | SS-L | -0.06 | PLIC-R | 0.01 | SLF-L | 0.01 | SCR-L | -0.22 |
| 11 | SS-L | -0.27 | CST-R | -0.29 | MCP | -0.06 | SFO-L | 0.01 | RLIC-L | 0.03 | CGC-R | -0.21 |
| 12 | RLIC-L | -0.26 | PCR-R | -0.28 | CGC-L | -0.05 | PLIC-L | 0.02 | PLIC-L | 0.03 | SLF-L | -0.21 |
| 13 | RLIC-R | -0.26 | FX/ST-R | -0.26 | CGH-R | -0.02 | ICP-L | 0.03 | ICP-L | 0.04 | RLIC-L | -0.20 |
| 14 | ACR-L | -0.25 | RLIC-L | -0.26 | SLF-L | -0.01 | MCP | 0.06 | ML-R | 0.07 | CGC-L | -0.20 |
| 15 | BCC | -0.24 | SLF-L | -0.26 | SCP-R | -0.01 | PTR-L | 0.06 | CGC-L | 0.07 | FX/ST-R | -0.20 |
| 16 | SCR-L | -0.23 | GCC | -0.24 | TAP-L | 0.00 | SCP-L | 0.09 | EC-L | 0.07 | SCR-R | -0.18 |
| 17 | UNC-R | -0.23 | TAP-R | -0.23 | TAP-R | 0.00 | CP-L | 0.09 | SFO-R | 0.08 | PCT | -0.18 |
| 18 | UNC-L | -0.22 | UNC-R | -0.23 | SCC | 0.01 | CST-L | 0.09 | PCT | 0.09 | ACR-R | -0.17 |
| 19 | SLF-R | -0.21 | SLF-R | -0.23 | PCT | 0.01 | CP-R | 0.09 | MCP | 0.10 | SCP-R | -0.16 |
| 20 | CGC-R | -0.18 | IFO-R | -0.22 | PCR-L | 0.02 | ACR-L | 0.11 | SFO-L | 0.10 | SCP-L | -0.16 |
| 21 | GCC | -0.17 | SCR-L | -0.19 | CST-R | 0.02 | SCC | 0.12 | IFO-L | 0.10 | SCC | -0.15 |
| 22 | SCR-R | -0.17 | ACR-L | -0.18 | SFO-R | 0.02 | SCR-L | 0.13 | CST-L | 0.11 | TAP-L | -0.15 |
| 23 | FX/ST-R | -0.15 | UNC-L | -0.18 | SLF-R | 0.03 | SS-L | 0.14 | TAP-R | 0.13 | CGH-L | -0.14 |
| 24 | SFO-R | -0.15 | ML-L | -0.18 | PLIC-L | 0.03 | PTR-R | 0.14 | ALIC-R | 0.14 | UNC-R | -0.14 |
| 25 | MCP | -0.13 | SFO-R | -0.16 | SCR-L | 0.04 | IFO-R | 0.15 | PCR-L | 0.16 | CST-R | -0.14 |
| 26 | TAP-L | -0.09 | FX/ST-L | -0.15 | CGC-R | 0.04 | TAP-R | 0.18 | FX/ST-L | 0.17 | MCP | -0.13 |
| 27 | PTR-L | -0.08 | PCR-L | -0.15 | CGH-L | 0.04 | CGH-L | 0.18 | ICP-R | 0.17 | CGH-R | -0.13 |
| 28 | PCT | -0.08 | MCP | -0.14 | FX/ST-L | 0.04 | CST-R | 0.18 | IFO-R | 0.18 | UNC-L | -0.13 |
| 29 | CGH-R | -0.08 | RLIC-R | -0.14 | ACR-L | 0.05 | SCR-R | 0.19 | CGC-R | 0.18 | SFO-R | -0.12 |
| 30 | IFO-R | -0.06 | SCR-R | -0.13 | SS-R | 0.06 | SFO-R | 0.19 | RLIC-R | 0.18 | GCC | -0.12 |
| 31 | SFO-L | -0.04 | CP-L | -0.12 | ICP-R | 0.06 | FX/ST-L | 0.21 | PTR-R | 0.19 | ICP-L | -0.12 |
| 32 | ICP-L | -0.01 | CP-R | -0.08 | PLIC-R | 0.06 | SCP-R | 0.21 | ML-L | 0.19 | PCR-L | -0.11 |
| 33 | PLIC-R | 0.00 | CGH-R | -0.07 | ALIC-L | 0.06 | ML-L | 0.23 | FX/ST-R | 0.20 | IFO-R | -0.11 |
| 34 | ML-R | 0.01 | EC-L | -0.07 | SCP-L | 0.07 | RLIC-L | 0.24 | CP-L | 0.26 | EC-L | -0.11 |
| 35 | CGH-L | 0.01 | CST-L | -0.07 | SFO-L | 0.07 | CGC-R | 0.25 | CGH-R | 0.26 | PTR-L | -0.09 |
| 36 | FX/ST-L | 0.01 | SCC | -0.07 | ALIC-R | 0.08 | PCR-L | 0.25 | CGH-L | 0.26 | ALIC-L | -0.09 |
| 37 | ML-L | 0.02 | TAP-L | -0.05 | IFO-L | 0.10 | CGC-L | 0.26 | SS-R | 0.28 | TAP-R | -0.09 |
| 38 | CST-R | 0.02 | ALIC-R | -0.05 | ML-L | 0.13 | SLF-L | 0.26 | PLIC-R | 0.29 | FX/ST-L | -0.08 |
| 39 | SCC | 0.02 | PTR-L | -0.01 | IFO-R | 0.15 | UNC-R | 0.30 | SCR-R | 0.29 | PTR-R | -0.07 |
| 40 | SCP-L | 0.03 | SCP-R | -0.00 | RLIC-L | 0.16 | EC-R | 0.32 | PCR-R | 0.30 | ML-L | -0.04 |
| 41 | CST-L | 0.04 | ALIC-L | 0.00 | PTR-L | 0.20 | SLF-R | 0.32 | ACR-L | 0.30 | SS-R | -0.04 |
| 42 | IFO-L | 0.07 | IFO-L | 0.00 | GCC | 0.20 | ML-R | 0.33 | GCC | 0.32 | CP-R | -0.02 |
| 43 | ICP-R | 0.08 | ICP-L | 0.02 | CP-R | 0.21 | CGH-R | 0.34 | CP-R | 0.33 | IFO-L | -0.00 |
| 44 | ALIC-L | 0.09 | ICP-R | 0.05 | PCR-R | 0.22 | FX/ST-R | 0.35 | PTR-L | 0.33 | ICP-R | 0.01 |
| 45 | EC-L | 0.10 | SCP-L | 0.05 | BCC | 0.26 | RLIC-R | 0.35 | SCP-R | 0.37 | CST-L | 0.02 |
| 46 | PLIC-L | 0.11 | PCT | 0.06 | RLIC-R | 0.28 | ACR-R | 0.45 | ACR-R | 0.37 | PLIC-L | 0.02 |
| 47 | SCP-R | 0.15 | PLIC-L | 0.06 | ACR-R | 0.28 | GCC | 0.47 | SCP-L | 0.39 | ML-R | 0.03 |
| 48 | CP-R | 0.17 | CGH-L | 0.08 | CST-L | 0.29 | BCC | 0.48 | SLF-R | 0.42 | SFO-L | 0.05 |
| 49 | CP-L | 0.21 | SFO-L | 0.09 | ML-R | 0.30 | SS-R | 0.52 | BCC | 0.43 | ALIC-R | 0.06 |
| 50 | ALIC-R | 0.22 | PLIC-R | 0.16 | SCR-R | 0.31 | PCR-R | 0.53 | SCC | 0.57 | RLIC-R | 0.06 |

| | $rd-SZ$ | | $md-SZ$ | | $ad-SZ$ | | $fa-SZ$ | | $rk-SZ$ | | $mk-SZ$ | |
|---|---|---|---|---|---|---|---|---|---|---|---|---|
| 1 | PCT | -0.21 | PCT | -0.27 | PCT | -0.28 | SFO-R | -0.49 | SCR-L | -0.67 | SCR-L | -0.70 |
| 2 | ICP-L | -0.06 | ICP-L | -0.14 | ICP-L | -0.22 | PTR-L | -0.41 | ALIC-L | -0.63 | SS-R | -0.65 |
| 3 | CST-L | -0.05 | ML-L | 0.01 | CP-L | -0.17 | ACR-R | -0.41 | ACR-L | -0.57 | SFO-L | -0.61 |
| 4 | CST-R | 0.01 | CST-R | 0.01 | ML-L | -0.13 | SCP-R | -0.39 | SS-R | -0.53 | SCR-R | -0.60 |
| 5 | PLIC-L | 0.03 | CST-L | 0.03 | ICP-R | -0.11 | ALIC-R | -0.35 | PTR-L | -0.50 | SLF-L | -0.60 |
| 6 | IFO-L | 0.08 | CP-L | 0.03 | CGC-R | -0.02 | ACR-L | -0.31 | SCR-R | -0.50 | TAP-R | -0.56 |
| 7 | EC-R | 0.09 | ICP-R | 0.06 | CST-R | 0.01 | SS-L | -0.30 | MCP | -0.49 | PTR-L | -0.56 |
| 8 | EC-L | 0.09 | EC-L | 0.08 | ALIC-R | 0.03 | CP-L | -0.29 | SLF-L | -0.48 | ACR-L | -0.55 |
| 9 | UNC-R | 0.10 | PLIC-L | 0.11 | SFO-R | 0.04 | CGH-R | -0.29 | TAP-R | -0.47 | PTR-R | -0.53 |
| 10 | IFO-R | 0.11 | EC-R | 0.14 | ML-R | 0.05 | FX/ST-L | -0.29 | FX | -0.42 | PCR-L | -0.50 |
| 11 | ML-L | 0.13 | IFO-L | 0.14 | CP-R | 0.06 | CGC-L | -0.29 | SS-L | -0.42 | FX | -0.48 |
| 12 | CP-L | 0.16 | CP-R | 0.15 | EC-L | 0.06 | FX | -0.28 | ACR-R | -0.38 | BCC | -0.48 |
| 13 | ICP-R | 0.16 | IFO-R | 0.20 | ALIC-L | 0.08 | SCR-R | -0.26 | PTR-R | -0.38 | SLF-R | -0.47 |
| 14 | PCR-L | 0.18 | ML-R | 0.20 | CGC-L | 0.09 | SS-R | -0.26 | SLF-R | -0.37 | ACR-R | -0.44 |
| 15 | UNC-L | 0.18 | ALIC-L | 0.20 | CST-L | 0.13 | CGC-R | -0.26 | CGC-L | -0.37 | SS-L | -0.43 |
| 16 | MCP | 0.20 | PCR-L | 0.20 | SS-L | 0.17 | SFO-L | -0.26 | BCC | -0.34 | PCR-R | -0.42 |
| 17 | CP-R | 0.20 | MCP | 0.21 | SCR-R | 0.17 | BCC | -0.25 | SFO-L | -0.34 | CGC-L | -0.42 |
| 18 | PLIC-R | 0.22 | UNC-L | 0.22 | EC-R | 0.19 | SCP-L | -0.24 | ALIC-R | -0.31 | MCP | -0.38 |
| 19 | ALIC-L | 0.25 | CGC-R | 0.23 | UNC-L | 0.19 | ML-R | -0.24 | CGC-R | -0.31 | TAP-L | -0.38 |
| 20 | SCR-L | 0.25 | UNC-R | 0.25 | SFO-L | 0.19 | SCC | -0.22 | FX/ST-R | -0.30 | SCC | -0.37 |
| 21 | RLIC-L | 0.25 | RLIC-R | 0.27 | IFO-L | 0.20 | RLIC-R | -0.21 | PCR-R | -0.29 | CGC-R | -0.36 |
| 22 | ML-R | 0.26 | RLIC-L | 0.27 | PCR-L | 0.21 | ICP-R | -0.20 | PCR-L | -0.29 | RLIC-R | -0.34 |
| 23 | RLIC-R | 0.26 | SS-L | 0.28 | PLIC-L | 0.22 | CP-R | -0.20 | ML-R | -0.28 | SFO-R | -0.32 |
| 24 | CGC-R | 0.29 | ALIC-R | 0.29 | MCP | 0.23 | RLIC-L | -0.19 | SFO-R | -0.26 | FX/ST-R | -0.29 |
| 25 | SLF-L | 0.30 | SCR-L | 0.30 | SS-R | 0.23 | GCC | -0.19 | EC-R | -0.26 | GCC | -0.28 |
| 26 | SS-L | 0.30 | SFO-L | 0.33 | RLIC-R | 0.25 | ML-L | -0.19 | ICP-R | -0.25 | PLIC-L | -0.25 |
| 27 | TAP-R | 0.30 | PLIC-R | 0.33 | RLIC-L | 0.25 | ALIC-L | -0.17 | TAP-L | -0.25 | EC-R | -0.25 |
| 28 | PTR-R | 0.33 | FX | 0.34 | IFO-R | 0.27 | SLF-R | -0.17 | GCC | -0.24 | RLIC-L | -0.22 |
| 29 | SCC | 0.33 | TAP-L | 0.34 | FX/ST-L | 0.27 | FX/ST-R | -0.16 | UNC-R | -0.22 | ICP-R | -0.21 |
| 30 | BCC | 0.33 | SCC | 0.35 | PTR-L | 0.28 | PTR-R | -0.16 | ICP-L | -0.21 | ALIC-L | -0.20 |
| 31 | TAP-L | 0.33 | CGC-L | 0.35 | PLIC-R | 0.29 | TAP-L | -0.15 | RLIC-L | -0.20 | SCP-L | -0.19 |
| 32 | FX | 0.34 | TAP-R | 0.36 | SCC | 0.29 | ICP-L | -0.14 | PLIC-L | -0.20 | ALIC-R | -0.18 |
| 33 | SLF-R | 0.35 | SLF-L | 0.36 | SCR-L | 0.30 | TAP-R | -0.14 | CP-L | -0.19 | SCP-R | -0.17 |
| 34 | GCC | 0.35 | SCP-L | 0.37 | SCP-L | 0.30 | PCR-R | -0.12 | RLIC-R | -0.17 | CST-L | -0.17 |
| 35 | SCP-L | 0.36 | SCR-R | 0.37 | PTR-R | 0.30 | SCR-L | -0.11 | FX/ST-L | -0.15 | ICP-L | -0.17 |
| 36 | CGH-L | 0.37 | PTR-R | 0.37 | TAP-L | 0.31 | SLF-L | -0.10 | IFO-R | -0.13 | ML-R | -0.14 |
| 37 | PCR-R | 0.37 | CGH-L | 0.38 | FX | 0.31 | MCP | -0.09 | ML-L | -0.11 | ML-L | -0.12 |
| 38 | CGC-L | 0.39 | BCC | 0.38 | SCP-R | 0.32 | CGH-L | -0.09 | SCC | -0.11 | FX/ST-L | -0.10 |
| 39 | ALIC-R | 0.39 | FX/ST-L | 0.39 | ACR-L | 0.33 | PCR-L | -0.02 | SCP-R | -0.10 | PLIC-R | -0.08 |
| 40 | SFO-L | 0.41 | SS-R | 0.42 | BCC | 0.34 | PLIC-R | -0.01 | CST-L | -0.09 | IFO-R | -0.08 |
| 41 | FX/ST-R | 0.43 | FX/ST-R | 0.43 | UNC-R | 0.34 | UNC-L | 0.01 | CST-R | -0.02 | EC-L | -0.05 |
| 42 | SCR-R | 0.43 | SFO-R | 0.44 | CGH-L | 0.34 | EC-L | 0.01 | EC-L | -0.01 | UNC-L | -0.02 |
| 43 | FX/ST-L | 0.44 | SLF-R | 0.44 | SLF-L | 0.38 | CST-R | 0.01 | SCP-L | -0.01 | CGH-L | -0.01 |
| 44 | SS-R | 0.46 | PCR-R | 0.45 | TAP-R | 0.41 | PCT | 0.01 | UNC-L | 0.00 | UNC-R | -0.00 |
| 45 | PTR-L | 0.47 | GCC | 0.45 | FX/ST-R | 0.41 | IFO-R | 0.04 | IFO-L | 0.01 | IFO-L | 0.03 |
| 46 | ACR-L | 0.48 | PTR-L | 0.45 | SLF-R | 0.42 | PLIC-L | 0.04 | PLIC-R | 0.03 | CGH-R | 0.05 |
| 47 | CGH-R | 0.53 | ACR-L | 0.48 | CGH-R | 0.42 | IFO-L | 0.06 | CP-R | 0.06 | CP-L | 0.05 |
| 48 | SCP-R | 0.53 | SCP-R | 0.49 | GCC | 0.44 | UNC-R | 0.08 | CGH-L | 0.15 | CST-R | 0.05 |
| 49 | SFO-R | 0.57 | CGH-R | 0.51 | PCR-R | 0.44 | CST-L | 0.09 | CGH-R | 0.17 | CP-R | 0.09 |
| 50 | ACR-R | 0.63 | ACR-R | 0.69 | ACR-R | 0.48 | EC-R | 0.11 | PCT | 0.23 | PCT | 0.22 |

| | $ak-SZ$ | | $f-SZ$ | | $da-SZ$ | | $depar-SZ$ | | $deperp-SZ$ | | $c2-SZ$ | |
|---|---|---|---|---|---|---|---|---|---|---|---|---|
| 1 | SCR-L | -0.84 | SCR-L | -0.69 | FX | -0.35 | PCT | -0.36 | ALIC-L | -0.30 | ACR-R | -0.43 |
| 2 | SLF-L | -0.61 | ALIC-L | -0.59 | PCT | -0.26 | FX | -0.35 | SCR-L | -0.28 | CP-L | -0.34 |
| 3 | SLF-R | -0.56 | ACR-L | -0.48 | TAP-L | -0.23 | TAP-L | -0.24 | FX | -0.17 | FX/ST-L | -0.32 |
| 4 | BCC | -0.49 | SLF-L | -0.42 | FX/ST-R | -0.15 | CST-L | -0.23 | TAP-L | -0.17 | ALIC-L | -0.29 |
| 5 | TAP-R | -0.49 | FX | -0.41 | UNC-R | -0.13 | CP-R | -0.22 | UNC-R | -0.14 | ALIC-R | -0.28 |
| 6 | SCR-R | -0.46 | SS-L | -0.40 | FX/ST-L | -0.12 | CST-R | -0.18 | PCT | -0.12 | FX | -0.28 |
| 7 | PTR-R | -0.45 | BCC | -0.37 | SCP-R | -0.08 | SCC | -0.15 | ICP-L | -0.11 | TAP-L | -0.27 |
| 8 | ACR-R | -0.43 | PTR-R | -0.34 | MCP | -0.01 | CGH-L | -0.11 | PLIC-L | -0.08 | ACR-L | -0.24 |
| 9 | ACR-L | -0.43 | FX/ST-R | -0.34 | TAP-R | -0.01 | PLIC-R | -0.09 | CST-L | -0.08 | ML-R | -0.23 |
| 10 | SFO-L | -0.42 | ACR-R | -0.34 | ML-L | 0.00 | EC-L | -0.06 | CP-L | -0.06 | SFO-R | -0.22 |
| 11 | PCR-R | -0.40 | PTR-L | -0.33 | ICP-L | 0.00 | ICP-L | -0.03 | IFO-R | -0.03 | BCC | -0.20 |
| 12 | PTR-L | -0.38 | SCR-R | -0.33 | CP-L | 0.02 | CP-L | -0.02 | CST-R | -0.01 | PTR-L | -0.19 |
| 13 | PCR-L | -0.37 | CGC-L | -0.33 | SS-L | 0.04 | ML-L | -0.01 | SLF-L | -0.00 | ICP-R | -0.17 |
| 14 | SS-R | -0.36 | MCP | -0.32 | CST-R | 0.04 | ICP-R | 0.01 | EC-R | 0.02 | CGC-L | -0.16 |
| 15 | SS-L | -0.31 | CP-L | -0.32 | SFO-L | 0.05 | UNC-L | 0.02 | SCR-R | 0.03 | SS-L | -0.16 |
| 16 | FX | -0.31 | SS-R | -0.31 | SCP-L | 0.06 | SFO-R | 0.03 | MCP | 0.04 | SCP-R | -0.15 |
| 17 | CGC-L | -0.30 | SLF-R | -0.30 | ALIC-R | 0.07 | SCP-L | 0.05 | IFO-L | 0.04 | SFO-L | -0.14 |
| 18 | TAP-L | -0.28 | ALIC-R | -0.27 | SCC | 0.10 | RLIC-R | 0.07 | CGC-R | 0.05 | CGH-R | -0.13 |
| 19 | CGC-R | -0.23 | UNC-R | -0.26 | ICP-R | 0.12 | EC-R | 0.08 | ML-L | 0.05 | CGC-R | -0.13 |
| 20 | SCC | -0.22 | CGC-R | -0.26 | CGC-L | 0.12 | IFO-L | 0.08 | ICP-R | 0.06 | ML-L | -0.12 |
| 21 | MCP | -0.20 | ICP-L | -0.25 | PTR-L | 0.13 | PCR-L | 0.08 | SS-L | 0.11 | FX/ST-R | -0.11 |
| 22 | PLIC-L | -0.20 | PLIC-L | -0.25 | RLIC-L | 0.14 | CGH-R | 0.12 | ALIC-R | 0.12 | PCT | -0.11 |
| 23 | UNC-R | -0.17 | ML-R | -0.24 | ML-R | 0.15 | MCP | 0.13 | SLF-R | 0.13 | UNC-R | -0.11 |
| 24 | ALIC-L | -0.17 | RLIC-L | -0.24 | SFO-R | 0.17 | ML-R | 0.15 | ML-R | 0.14 | SCR-R | -0.09 |
| 25 | FX/ST-R | -0.16 | FX/ST-L | -0.22 | CGC-R | 0.17 | SCP-R | 0.16 | RLIC-L | 0.14 | RLIC-L | -0.08 |
| 26 | RLIC-L | -0.11 | EC-R | -0.20 | CGH-R | 0.19 | IFO-R | 0.18 | EC-L | 0.16 | GCC | -0.08 |
| 27 | IFO-R | -0.08 | PCR-R | -0.20 | ACR-L | 0.20 | SFO-L | 0.18 | CGC-L | 0.19 | CP-R | -0.06 |
| 28 | EC-R | -0.03 | TAP-R | -0.20 | PTR-R | 0.20 | ALIC-R | 0.24 | SFO-L | 0.22 | SCR-L | -0.06 |
| 29 | FX/ST-L | -0.03 | TAP-L | -0.19 | ALIC-L | 0.21 | FX/ST-L | 0.25 | UNC-L | 0.23 | MCP | -0.06 |
| 30 | ALIC-R | 0.00 | ICP-R | -0.16 | UNC-L | 0.22 | CGC-R | 0.25 | BCC | 0.23 | CST-R | -0.06 |
| 31 | RLIC-R | 0.04 | IFO-R | -0.15 | EC-R | 0.24 | GCC | 0.27 | ACR-L | 0.25 | CGH-L | -0.06 |
| 32 | UNC-L | 0.05 | SFO-L | -0.13 | IFO-R | 0.25 | PCR-R | 0.28 | PCR-L | 0.26 | SS-R | -0.05 |
| 33 | IFO-L | 0.05 | ML-L | -0.09 | ACR-R | 0.26 | SS-R | 0.28 | PCR-R | 0.26 | SLF-R | -0.03 |
| 34 | GCC | 0.07 | SFO-R | -0.07 | IFO-L | 0.26 | PTR-R | 0.29 | FX/ST-R | 0.28 | ICP-L | -0.01 |
| 35 | ML-R | 0.08 | RLIC-R | -0.07 | PLIC-L | 0.26 | TAP-R | 0.30 | RLIC-R | 0.29 | SLF-L | -0.01 |
| 36 | CST-L | 0.08 | PCR-L | -0.07 | SCR-L | 0.27 | SS-L | 0.30 | SS-R | 0.29 | UNC-L | 0.00 |
| 37 | SFO-R | 0.08 | CST-R | -0.03 | CST-L | 0.28 | PLIC-L | 0.32 | FX/ST-L | 0.29 | IFO-R | -0.00 |
| 38 | SCP-R | 0.12 | UNC-L | -0.02 | BCC | 0.30 | RLIC-L | 0.33 | PLIC-R | 0.30 | SCC | 0.01 |
| 39 | CST-R | 0.14 | GCC | -0.02 | RLIC-R | 0.31 | SCR-R | 0.33 | PTR-R | 0.31 | IFO-L | 0.01 |
| 40 | CGH-L | 0.15 | CST-L | 0.00 | SS-R | 0.31 | FX/ST-R | 0.34 | SFO-R | 0.33 | PCR-R | 0.02 |
| 41 | SCP-L | 0.16 | IFO-L | -0.00 | SLF-L | 0.32 | CGC-L | 0.36 | ACR-R | 0.34 | TAP-R | 0.05 |
| 42 | PLIC-R | 0.18 | PCT | 0.05 | SLF-R | 0.33 | SLF-R | 0.38 | TAP-R | 0.35 | CST-L | 0.07 |
| 43 | CGH-R | 0.19 | SCC | 0.06 | CP-R | 0.34 | SLF-L | 0.40 | CP-R | 0.36 | RLIC-R | 0.11 |
| 44 | ML-L | 0.19 | PLIC-R | 0.08 | EC-L | 0.36 | PTR-L | 0.40 | SCC | 0.36 | PTR-R | 0.11 |
| 45 | ICP-L | 0.20 | CP-R | 0.09 | PCR-R | 0.39 | BCC | 0.45 | GCC | 0.38 | PLIC-L | 0.14 |
| 46 | PCT | 0.22 | SCP-R | 0.10 | PCR-L | 0.40 | UNC-R | 0.48 | CGH-L | 0.39 | SCP-L | 0.15 |
| 47 | ICP-R | 0.24 | EC-L | 0.11 | PLIC-R | 0.41 | ACR-L | 0.50 | PTR-L | 0.40 | EC-L | 0.17 |
| 48 | EC-L | 0.26 | CGH-R | 0.13 | CGH-L | 0.41 | ACR-R | 0.65 | SCP-R | 0.56 | PCR-L | 0.17 |
| 49 | CP-L | 0.31 | CGH-L | 0.20 | SCR-R | 0.42 | ALIC-L | 0.68 | CGH-R | 0.57 | PLIC-R | 0.19 |
| 50 | CP-R | 0.36 | SCP-L | 0.28 | GCC | 0.62 | SCR-L | 0.78 | SCP-L | 0.63 | EC-R | 0.22 |

Table S 4: Sorted effect sizes for EP and SZ for each dMRI metric and ROI

| | metric | RD | MD | AD | FA | RK | MK | AK | $f$ | $D_a$ | $D_{e,\parallel}$ | $D_{\perp}$ | $c_2$ |
|---|---|---|---|---|---|---|---|---|---|---|---|---|---|
| 1 | MIDCP | 0.2437, 0.28, [0.01, 0.55] | 0.2008, 0.31, [0.04, 0.58] | 0.1195, 0.35, [0.08, 0.62] | 0.475, -0.05, [-0.32, 0.22] | 0.2472, -0.13, [-0.4, 0.14] | 0.2396, -0.15, [-0.42, 0.12] | 0.3298, -0.13, [-0.4, 0.14] | 0.4234, -0.14, [-0.4, 0.13] | 0.3298, -0.06, [-0.32, 0.21] | 0.3624, 0.06, [-0.21, 0.33] | 0.4053, 0.1, [-0.16, 0.37] | 0.2229, -0.13, [-0.4, 0.13] |
| 2 | PCT | 0.4645, 0.05, [-0.22, 0.32] | 0.4557, 0.05, [-0.22, 0.32] | 0.4443, 0.03, [-0.24, 0.3] | 0.3552, -0.07, [-0.34, 0.19] | 0.1389, 0.22, [-0.05, 0.49] | 0.3372, 0.14, [-0.13, 0.41] | 0.2039, -0.08, [-0.35, 0.19] | 0.3023, 0.06, [-0.2, 0.33] | 0.4506, 0.01, [-0.26, 0.28] | 0.4511, -0.01, [-0.27, 0.26] | 0.2407, 0.09, [-0.18, 0.36] | 0.2075, -0.18, [-0.45, 0.08] |
| 3 | GCC | **0.0085**, 0.38, [0.11, 0.65] | **0.0027**, 0.48, [0.21, 0.75] | **0.009**, 0.39, [0.12, 0.65] | 0.0746, -0.19, [-0.46, 0.07] | **0.0282**, -0.3, [-0.57, -0.03] | **0.0238**, -0.32, [-0.59, -0.05] | 0.1631, -0.17, [-0.44, 0.1] | 0.0583, -0.24, [-0.51, 0.03] | 0.1117, 0.2, [-0.07, 0.47] | **0.0041**, 0.47, [0.2, 0.74] | **0.0344**, 0.32, [0.05, 0.58] | 0.1882, -0.12, [-0.38, 0.15] |
| 4 | BCC | **0.0023**, 0.51, [0.24, 0.78] | **0.0002**, 0.54, [0.27, 0.81] | **0.0085**, 0.37, [0.1, 0.63] | **0.014**, -0.34, [-0.61, -0.08] | **0.0178**, -0.32, [-0.59, -0.06] | **0.0044**, -0.41, [-0.68, -0.14] | 0.0583, -0.24, [-0.51, 0.03] | **0.0238**, -0.3, [-0.57, -0.03] | 0.0576, 0.26, [-0.0, 0.53] | **0.0017**, 0.48, [0.21, 0.75] | **0.0044**, 0.43, [0.16, 0.7] | **0.0258**, -0.32, [-0.59, -0.05] |
| 5 | SCC | **0.0021**, 0.55, [0.28, 0.82] | **0.0044**, 0.53, [0.26, 0.8] | 0.1148, 0.27, [0.0, 0.54] | **0.0044**, -0.37, [-0.63, -0.1] | 0.1183, -0.22, [-0.49, 0.05] | **0.0443**, -0.29, [-0.56, -0.02] | 0.4376, 0.02, [-0.25, 0.28] | 0.2868, -0.07, [-0.33, 0.2] | 0.4264, 0.01, [-0.26, 0.28] | 0.3553, 0.12, [-0.15, 0.38] | **0.002**, 0.57, [0.3, 0.84] | 0.0791, -0.15, [-0.42, 0.12] |
| 6 | FX | **0.0021**, 0.49, [0.22, 0.76] | **0.003**, 0.5, [0.22, 0.76] | **0.0248**, 0.43, [0.16, 0.7] | **0.033**, -0.33, [-0.6, -0.06] | 0.057, -0.26, [-0.53, 0.01] | **0.0124**, -0.37, [-0.64, -0.1] | 0.0639, -0.28, [-0.55, -0.01] | **0.0045**, -0.44, [-0.71, -0.17] | **0.036**, -0.29, [-0.55, -0.02] | 0.1466, -0.2, [-0.47, 0.07] | 0.4669, 0.01, [-0.26, 0.27] | 0.0639, -0.27, [-0.54, -0.0] |
| 7 | CST R | 0.151, 0.23, [-0.04, 0.5] | 0.1892, 0.22, [-0.05, 0.49] | 0.22, 0.16, [-0.11, 0.42] | 0.3065, -0.07, [-0.34, 0.2] | 0.0563, -0.28, [-0.55, -0.02] | 0.097, -0.15, [-0.42, 0.11] | 0.4842, 0.02, [-0.25, 0.29] | 0.0518, -0.29, [-0.56, -0.02] | 0.3394, 0.02, [-0.24, 0.29] | 0.1058, 0.18, [-0.09, 0.45] | 0.1822, -0.13, [-0.4, 0.13] | 0.4598, -0.14, [-0.41, 0.13] |
| 8 | CST L | 0.241, 0.25, [-0.02, 0.51] | 0.0994, 0.31, [0.04, 0.58] | 0.0669, 0.34, [0.07, 0.61] | 0.38, -0.02, [-0.29, 0.24] | 0.3053, -0.09, [-0.36, 0.18] | 0.2742, -0.1, [-0.36, 0.17] | 0.3976, 0.04, [-0.22, 0.31] | 0.4458, -0.07, [-0.34, 0.2] | **0.0238**, 0.29, [0.02, 0.56] | 0.4654, 0.09, [-0.17, 0.36] | 0.2982, 0.11, [-0.15, 0.38] | 0.2894, 0.02, [-0.25, 0.29] |
| 9 | ML R | 0.1419, 0.2, [-0.08, 0.47] | 0.2067, 0.15, [-0.12, 0.42] | 0.3159, 0.06, [-0.21, 0.33] | 0.255, -0.16, [-0.43, 0.11] | **0.014**, -0.34, [-0.62, -0.07] | **0.033**, -0.25, [-0.52, 0.03] | 0.449, 0.01, [-0.27, 0.28] | **0.014**, -0.3, [-0.57, -0.02] | 0.1747, 0.3, [0.02, 0.57] | 0.083, 0.33, [0.05, 0.6] | 0.4234, 0.07, [-0.2, 0.34] | 0.3723, 0.03, [-0.24, 0.3] |
| 10 | ML L | 0.1945, 0.18, [-0.09, 0.45] | 0.3073, 0.13, [-0.14, 0.4] | 0.4682, 0.07, [-0.21, 0.34] | 0.314, -0.15, [-0.42, 0.12] | 0.0639, -0.26, [-0.53, 0.01] | 0.0576, -0.2, [-0.47, 0.07] | 0.38, 0.02, [-0.25, 0.3] | 0.1524, -0.18, [-0.45, 0.09] | 0.38, 0.13, [-0.14, 0.4] | 0.1631, 0.23, [-0.04, 0.5] | 0.2113, 0.19, [-0.08, 0.46] | 0.4155, -0.04, [-0.31, 0.23] |
| 11 | ICP R | 0.4654, 0.0, [-0.28, 0.28] | 0.4487, -0.01, [-0.28, 0.27] | 0.3919, -0.03, [-0.3, 0.25] | 0.2439, -0.1, [-0.38, 0.18] | 0.4871, -0.01, [-0.29, 0.27] | 0.3336, -0.05, [-0.33, 0.22] | 0.2982, 0.08, [-0.2, 0.36] | 0.38, 0.05, [-0.23, 0.33] | 0.4616, 0.06, [-0.22, 0.33] | 0.4669, -0.02, [-0.29, 0.26] | 0.2103, 0.17, [-0.11, 0.45] | 0.4116, 0.01, [-0.27, 0.29] |
| 12 | ICP L | 0.474, 0.12, [-0.16, 0.39] | 0.3496, 0.08, [-0.2, 0.36] | 0.3655, -0.01, [-0.29, 0.27] | 0.4264, -0.12, [-0.4, 0.16] | 0.4193, 0.03, [-0.25, 0.31] | 0.4378, 0.0, [-0.27, 0.28] | 0.475, -0.01, [-0.29, 0.27] | 0.385, 0.02, [-0.26, 0.29] | 0.3344, -0.19, [-0.46, 0.09] | 0.4901, 0.03, [-0.25, 0.3] | 0.4531, 0.04, [-0.24, 0.31] | 0.4895, -0.12, [-0.4, 0.16] |
| 13 | SCP R | **0.0143**, 0.4, [0.13, 0.67] | **0.0216**, 0.38, [0.11, 0.65] | 0.0539, 0.27, [0.0, 0.54] | 0.0743, -0.27, [-0.54, 0.0] | 0.2892, -0.12, [-0.38, 0.15] | 0.1445, -0.07, [-0.34, 0.2] | 0.2301, 0.15, [-0.12, 0.41] | 0.4834, -0.0, [-0.27, 0.27] | 0.4414, -0.01, [-0.27, 0.26] | 0.1238, 0.21, [-0.06, 0.48] | **0.0275**, 0.37, [0.1, 0.64] | 0.1899, -0.16, [-0.43, 0.11] |
| 14 | SCP L | **0.0251**, 0.39, [0.12, 0.66] | **0.0369**, 0.39, [0.12, 0.66] | 0.1253, 0.29, [0.02, 0.56] | 0.0755, -0.28, [-0.54, -0.01] | 0.3298, -0.09, [-0.35, 0.18] | 0.0961, -0.2, [-0.47, 0.07] | 0.4789, 0.03, [-0.24, 0.3] | 0.4116, 0.05, [-0.22, 0.32] | 0.212, 0.07, [-0.2, 0.34] | 0.2845, 0.09, [-0.17, 0.36] | **0.0181**, 0.39, [0.12, 0.66] | 0.3726, -0.16, [-0.43, 0.11] |
| 15 | CP R | **0.0153**, 0.38, [0.11, 0.65] | **0.0133**, 0.37, [0.1, 0.64] | **0.0467**, 0.29, [0.03, 0.56] | 0.0846, -0.18, [-0.45, 0.08] | 0.214, -0.17, [-0.44, 0.09] | 0.2301, -0.11, [-0.38, 0.16] | 0.1875, 0.17, [-0.1, 0.43] | 0.4475, -0.08, [-0.35, 0.19] | 0.0846, 0.21, [-0.06, 0.47] | 0.2723, 0.09, [-0.17, 0.36] | 0.056, 0.33, [0.06, 0.6] | 0.4458, -0.02, [-0.29, 0.25] |
| 16 | CP L | **0.0063**, 0.48, [0.21, 0.75] | 0.0639, 0.32, [0.05, 0.59] | 0.4156, 0.04, [-0.23, 0.3] | **0.0002**, -0.4, [-0.67, -0.13] | 0.3924, -0.09, [-0.35, 0.18] | 0.4616, -0.01, [-0.28, 0.25] | 0.212, 0.21, [-0.05, 0.48] | 0.2896, -0.12, [-0.38, 0.15] | 0.126, -0.16, [-0.43, 0.1] | 0.2206, 0.09, [-0.18, 0.35] | 0.1249, 0.26, [-0.01, 0.53] | **0.014**, -0.31, [-0.58, -0.04] |
| 17 | ALIC R | 0.1536, 0.2, [-0.07, 0.46] | 0.2301, 0.12, [-0.14, 0.39] | 0.4006, -0.02, [-0.28, 0.25] | 0.1892, -0.17, [-0.44, 0.09] | 0.4268, -0.06, [-0.33, 0.21] | 0.3023, -0.1, [-0.36, 0.17] | 0.148, 0.22, [-0.05, 0.48] | 0.4496, -0.05, [-0.32, 0.22] | 0.3414, 0.08, [-0.18, 0.35] | 0.4834, -0.05, [-0.32, 0.21] | 0.2553, 0.14, [-0.13, 0.41] | 0.4165, 0.06, [-0.2, 0.33] |
| 18 | ALIC L | 0.174, -0.0, [-0.27, 0.27] | 0.2206, -0.04, [-0.3, 0.23] | 0.4979, -0.1, [-0.36, 0.17] | 0.1677, -0.11, [-0.37, 0.16] | 0.4881, -0.03, [-0.29, 0.24] | 0.4495, 0.04, [-0.22, 0.31] | 0.3187, 0.09, [-0.18, 0.36] | 0.4695, 0.0, [-0.27, 0.27] | 0.3185, 0.06, [-0.21, 0.32] | 0.2985, 0.0, [-0.26, 0.27] | 0.2075, -0.02, [-0.29, 0.25] | 0.2401, -0.09, [-0.36, 0.18] |
| 19 | PLIC R | 0.0639, 0.32, [0.06, 0.59] | 0.0746, 0.29, [0.02, 0.56] | 0.3024, 0.12, [-0.15, 0.39] | 0.0518, -0.29, [-0.56, -0.02] | 0.1191, 0.2, [-0.07, 0.47] | 0.3543, 0.05, [-0.22, 0.31] | 0.3976, 0.0, [-0.27, 0.27] | 0.2949, 0.16, [-0.1, 0.43] | 0.4492, 0.06, [-0.21, 0.32] | 0.4868, 0.01, [-0.25, 0.28] | 0.1117, 0.29, [0.02, 0.56] | 0.1011, -0.27, [-0.54, -0.01] |
| 20 | PLIC L | 0.2229, -0.04, [-0.3, 0.23] | 0.2694, -0.06, [-0.33, 0.2] | 0.3923, -0.09, [-0.35, 0.18] | 0.174, -0.05, [-0.32, 0.22] | 0.3298, 0.04, [-0.22, 0.31] | 0.4449, 0.03, [-0.24, 0.29] | 0.2537, 0.11, [-0.16, 0.37] | 0.358, 0.06, [-0.21, 0.32] | 0.4168, 0.03, [-0.23, 0.3] | 0.4669, 0.02, [-0.25, 0.28] | 0.1369, 0.03, [-0.24, 0.29] | 0.3951, 0.02, [-0.25, 0.28] |
| 21 | RPIC R | 0.1253, 0.06, [-0.2, 0.33] | **0.0313**, 0.1, [-0.17, 0.36] | 0.0825, 0.13, [-0.14, 0.4] | 0.2659, -0.09, [-0.36, 0.17] | 0.2145, -0.15, [-0.41, 0.12] | 0.0541, -0.3, [-0.57, -0.03] | 0.0583, -0.26, [-0.52, 0.01] | 0.2191, -0.14, [-0.41, 0.12] | 0.1143, 0.28, [0.02, 0.55] | **0.0248**, 0.35, [0.09, 0.62] | 0.2194, 0.18, [-0.09, 0.45] | 0.4834, 0.06, [-0.21, 0.32] |
| 22 | RPIC L | **0.0051**, 0.12, [-0.14, 0.39] | **0.0054**, 0.09, [-0.17, 0.36] | 0.2421, -0.0, [-0.27, 0.27] | **0.0164**, -0.27, [-0.54, -0.01] | 0.0804, -0.23, [-0.5, 0.04] | **0.0355**, -0.29, [-0.55, -0.02] | **0.042**, -0.26, [-0.52, 0.01] | 0.0949, -0.26, [-0.53, 0.0] | 0.1875, 0.16, [-0.11, 0.43] | **0.0045**, 0.24, [-0.03, 0.51] | 0.2851, 0.03, [-0.24, 0.3] | 0.212, -0.2, [-0.46, 0.07] |
| 23 | ACR R | **0.0008**, 0.58, [0.31, 0.85] | **0.0002**, 0.65, [0.38, 0.93] | **0.0054**, 0.45, [0.18, 0.71] | **0.0378**, -0.32, [-0.59, -0.05] | **0.003**, -0.51, [-0.78, -0.24] | **0.0017**, -0.56, [-0.84, -0.29] | **0.0271**, -0.39, [-0.65, -0.12] | **0.0155**, -0.36, [-0.63, -0.09] | 0.0945, 0.28, [0.01, 0.54] | **0.0045**, 0.45, [0.18, 0.72] | **0.0133**, 0.37, [0.1, 0.64] | 0.1222, -0.17, [-0.43, 0.1] |
| 24 | ACR L | **0.0045**, 0.26, [-0.01, 0.52] | **0.0021**, 0.22, [-0.05, 0.49] | 0.0576, 0.1, [-0.16, 0.37] | **0.0296**, -0.36, [-0.62, -0.09] | **0.0365**, -0.3, [-0.57, -0.04] | **0.007**, -0.41, [-0.68, -0.14] | 0.104, -0.25, [-0.52, 0.02] | 0.214, -0.18, [-0.45, 0.09] | 0.1892, 0.05, [-0.22, 0.32] | 0.0762, 0.11, [-0.15, 0.38] | **0.003**, 0.3, [0.03, 0.56] | 0.1044, -0.25, [-0.52, 0.02] |
| 25 | SCR R | **0.0024**, 0.5, [0.23, 0.77] | **0.0031**, 0.48, [0.21, 0.75] | 0.1483, 0.22, [-0.05, 0.48] | **0.0238**, -0.36, [-0.63, -0.09] | 0.0981, -0.29, [-0.56, -0.02] | 0.0909, -0.32, [-0.59, -0.06] | 0.1358, -0.17, [-0.44, 0.09] | 0.3298, -0.13, [-0.4, 0.14] | 0.0677, 0.31, [0.04, 0.57] | 0.2439, 0.19, [-0.08, 0.45] | 0.1049, 0.29, [0.02, 0.55] | 0.1875, -0.18, [-0.44, 0.09] |
| 26 | SCR L | **0.037**, 0.06, [-0.21, 0.32] | **0.0199**, 0.05, [-0.22, 0.31] | 0.152, -0.0, [-0.27, 0.27] | **0.0307**, -0.25, [-0.51, 0.02] | 0.1183, -0.22, [-0.48, 0.05] | 0.1038, -0.22, [-0.48, 0.05] | **0.0467**, -0.23, [-0.5, 0.04] | 0.1299, -0.19, [-0.45, 0.08] | 0.2553, 0.04, [-0.23, 0.3] | 0.0518, 0.13, [-0.14, 0.4] | 0.4616, -0.02, [-0.28, 0.25] | 0.1098, -0.22, [-0.49, 0.05] |
| 27 | PCR R | **0.0008**, 0.54, [0.26, 0.81] | **0.0002**, 0.61, [0.33, 0.88] | **0.0204**, 0.45, [0.18, 0.72] | **0.0164**, -0.31, [-0.57, -0.04] | **0.0378**, -0.35, [-0.62, -0.08] | **0.0102**, -0.45, [-0.72, -0.18] | **0.0092**, -0.46, [-0.73, -0.19] | 0.0669, -0.28, [-0.54, -0.01] | 0.2147, 0.22, [-0.05, 0.48] | **0.0019**, 0.53, [0.25, 0.8] | 0.0553, 0.3, [0.04, 0.57] | 0.0525, -0.27, [-0.54, -0.0] |
| 28 | PCR L | **0.0212**, 0.07, [-0.19, 0.34] | **0.0085**, 0.08, [-0.19, 0.34] | 0.0965, 0.06, [-0.21, 0.32] | 0.0755, -0.19, [-0.45, 0.08] | 0.0945, -0.22, [-0.48, 0.05] | **0.0138**, -0.35, [-0.61, -0.08] | **0.0286**, -0.4, [-0.67, -0.13] | 0.2271, -0.15, [-0.42, 0.12] | 0.4882, 0.02, [-0.25, 0.29] | 0.0583, 0.25, [-0.02, 0.52] | 0.0716, 0.16, [-0.11, 0.42] | 0.1475, -0.11, [-0.38, 0.16] |
| 29 | PTR R | 0.0518, 0.31, [0.04, 0.57] | **0.0478**, 0.29, [0.02, 0.56] | 0.1577, 0.18, [-0.09, 0.45] | 0.0583, -0.28, [-0.55, -0.01] | 0.0518, -0.34, [-0.61, -0.07] | **0.0114**, -0.46, [-0.73, -0.19] | 0.0945, -0.3, [-0.57, -0.03] | 0.1183, -0.3, [-0.56, -0.03] | 0.3228, -0.17, [-0.43, 0.1] | 0.2144, 0.14, [-0.12, 0.41] | 0.2226, 0.19, [-0.08, 0.46] | 0.4496, -0.07, [-0.34, 0.19] |
| 30 | PTR L | **0.0061**, 0.14, [-0.13, 0.41] | **0.0061**, 0.12, [-0.15, 0.39] | 0.1529, 0.05, [-0.22, 0.31] | **0.0107**, -0.32, [-0.59, -0.05] | 0.1195, -0.2, [-0.46, 0.07] | 0.0542, -0.27, [-0.54, -0.0] | 0.3503, -0.08, [-0.34, 0.19] | 0.475, -0.01, [-0.28, 0.25] | 0.2328, 0.2, [-0.07, 0.47] | 0.3632, 0.06, [-0.21, 0.32] | **0.0036**, 0.33, [0.07, 0.6] | 0.136, -0.09, [-0.36, 0.17] |
| 31 | SAGSTR R | **0.0043**, 0.42, [0.15, 0.68] | **0.0033**, 0.41, [0.14, 0.68] | 0.0639, 0.3, [0.03, 0.56] | **0.0248**, -0.29, [-0.56, -0.02] | **0.0044**, -0.45, [-0.72, -0.18] | **0.0019**, -0.54, [-0.81, -0.27] | **0.006**, -0.43, [-0.7, -0.16] | **0.0054**, -0.35, [-0.61, -0.08] | 0.4176, 0.06, [-0.21, 0.33] | **0.0017**, 0.52, [0.25, 0.79] | 0.0741, 0.28, [0.01, 0.54] | 0.3868, -0.04, [-0.31, 0.23] |
| 32 | SAGSTR L | **0.0133**, 0.19, [-0.08, 0.46] | **0.014**, 0.15, [-0.11, 0.42] | 0.1836, 0.04, [-0.23, 0.3] | **0.037**, -0.29, [-0.55, -0.02] | **0.014**, -0.38, [-0.65, -0.11] | **0.0102**, -0.42, [-0.69, -0.15] | 0.104, -0.27, [-0.54, -0.0] | **0.0378**, -0.33, [-0.6, -0.07] | 0.4592, -0.06, [-0.33, 0.21] | 0.097, 0.14, [-0.13, 0.41] | 0.3553, -0.03, [-0.3, 0.23] | 0.0974, -0.27, [-0.54, -0.0] |
| 33 | EC R | 0.0898, 0.2, [-0.07, 0.46] | 0.1697, 0.12, [-0.15, 0.39] | 0.475, -0.03, [-0.29, 0.24] | 0.0699, -0.28, [-0.55, -0.01] | 0.0576, -0.35, [-0.62, -0.08] | 0.0866, -0.27, [-0.53, 0.0] | **0.0344**, -0.32, [-0.59, -0.05] | **0.0378**, -0.37, [-0.63, -0.1] | 0.1295, -0.21, [-0.48, 0.06] | 0.0536, 0.32, [0.06, 0.59] | 0.4164, -0.09, [-0.35, 0.18] | 0.0775, -0.28, [-0.54, -0.01] |
| 34 | EC L | **0.0312**, 0.07, [-0.2, 0.34] | 0.0613, 0.01, [-0.25, 0.28] | 0.4032, -0.07, [-0.34, 0.19] | 0.1189, -0.24, [-0.51, 0.02] | 0.4669, -0.09, [-0.35, 0.18] | 0.4127, -0.05, [-0.32, 0.22] | 0.3298, 0.1, [-0.17, 0.37] | 0.4689, -0.07, [-0.34, 0.2] | 0.4557, -0.09, [-0.36, 0.18] | 0.3379, -0.04, [-0.3, 0.23] | 0.0511, 0.07, [-0.19, 0.34] | 0.4029, -0.11, [-0.37, 0.16] |
| 35 | CING/CG R | **0.0041**, 0.38, [0.11, 0.65] | **0.011**, 0.3, [0.03, 0.56] | 0.4627, 0.01, [-0.26, 0.27] | **0.0318**, -0.36, [-0.63, -0.09] | **0.0178**, -0.35, [-0.61, -0.08] | 0.0509, -0.31, [-0.57, -0.04] | 0.1067, -0.18, [-0.45, 0.08] | **0.0286**, -0.29, [-0.55, -0.02] | 0.3381, 0.04, [-0.23, 0.3] | **0.0251**, 0.25, [-0.02, 0.52] | 0.2039, 0.18, [-0.09, 0.45] | 0.0804, -0.21, [-0.48, 0.05] |
| 36 | CING/CG L | **0.0045**, 0.24, [-0.02, 0.51] | **0.0043**, 0.2, [-0.07, 0.46] | 0.4613, 0.0, [-0.26, 0.27] | **0.0493**, -0.26, [-0.53, 0.0] | **0.0378**, -0.3, [-0.57, -0.03] | 0.0673, -0.31, [-0.58, -0.04] | **0.0238**, -0.3, [-0.57, -0.04] | **0.0428**, -0.3, [-0.57, -0.04] | 0.361, -0.05, [-0.31, 0.22] | **0.005**, 0.26, [-0.01, 0.52] | 0.1892, 0.07, [-0.2, 0.34] | 0.0583, -0.2, [-0.47, 0.07] |
| 37 | CING/HIP R | **0.0054**, 0.43, [0.16, 0.7] | **0.0042**, 0.48, [0.21, 0.75] | **0.0116**, 0.43, [0.17, 0.7] | 0.2357, -0.11, [-0.38, 0.15] | 0.4116, -0.05, [-0.32, 0.21] | 0.474, -0.03, [-0.3, 0.24] | 0.3124, -0.08, [-0.35, 0.18] | 0.4164, -0.07, [-0.34, 0.2] | 0.4394, -0.02, [-0.28, 0.25] | 0.0547, 0.34, [0.07, 0.61] | **0.0369**, 0.26, [-0.01, 0.53] | 0.347, -0.13, [-0.39, 0.14] |
| 38 | CING/HIP L | **0.0104**, 0.42, [0.15, 0.69] | **0.0048**, 0.44, [0.17, 0.71] | **0.011**, 0.4, [0.13, 0.67] | 0.2463, -0.09, [-0.36, 0.18] | 0.1892, 0.18, [-0.09, 0.45] | 0.2609, 0.15, [-0.12, 0.41] | 0.4669, 0.01, [-0.26, 0.28] | 0.3061, 0.08, [-0.19, 0.34] | 0.3204, 0.04, [-0.23, 0.3] | 0.2017, 0.18, [-0.08, 0.45] | 0.0542, 0.26, [-0.01, 0.53] | 0.2526, -0.14, [-0.41, 0.12] |
| 39 | FX/STRIA R | **0.0175**, 0.37, [0.1, 0.64] | **0.024**, 0.36, [0.09, 0.62] | 0.0677, 0.31, [0.04, 0.57] | 0.0536, -0.31, [-0.58, -0.04] | 0.1186, -0.22, [-0.48, 0.05] | 0.1315, -0.22, [-0.49, 0.05] | 0.1735, -0.15, [-0.42, 0.11] | 0.0869, -0.26, [-0.52, 0.01] | 0.2917, -0.15, [-0.42, 0.11] | **0.0317**, 0.35, [0.08, 0.62] | 0.1515, 0.2, [-0.07, 0.47] | 0.1466, -0.2, [-0.46, 0.07] |
| 40 | FX/STRIA L | 0.0616, 0.12, [-0.14, 0.39] | 0.0969, 0.09, [-0.18, 0.35] | 0.3168, 0.02, [-0.25, 0.28] | 0.0677, -0.24, [-0.5, 0.03] | 0.1638, -0.19, [-0.46, 0.08] | 0.2342, -0.15, [-0.42, 0.12] | 0.4511, 0.01, [-0.25, 0.28] | 0.2379, -0.15, [-0.42, 0.11] | 0.4264, 0.04, [-0.23, 0.3] | 0.0559, 0.21, [-0.05, 0.48] | 0.1267, 0.17, [-0.1, 0.43] | 0.2896, -0.08, [-0.34, 0.19] |
| 41 | SLF R | **0.0001**, 0.57, [0.3, 0.84] | **0.0005**, 0.47, [0.2, 0.74] | 0.2518, 0.14, [-0.13, 0.4] | **0.0017**, -0.5, [-0.77, -0.23] | **0.0212**, -0.36, [-0.63, -0.1] | **0.0296**, -0.33, [-0.6, -0.06] | 0.0766, -0.21, [-0.48, 0.06] | 0.088, -0.23, [-0.5, 0.03] | 0.3804, 0.03, [-0.24, 0.3] | **0.0393**, 0.32, [0.05, 0.59] | **0.0048**, 0.42, [0.15, 0.69] | **0.0344**, -0.38, [-0.65, -0.11] |
| 42 | SLF L | **0.0044**, 0.18, [-0.09, 0.44] | **0.0049**, 0.12, [-0.15, 0.38] | 0.2891, -0.03, [-0.29, 0.24] | **0.023**, -0.31, [-0.58, -0.04] | **0.0124**, -0.4, [-0.67, -0.13] | **0.009**, -0.47, [-0.74, -0.2] | **0.006**, -0.43, [-0.7, -0.16] | 0.0551, -0.26, [-0.53, 0.0] | 0.4557, -0.01, [-0.28, 0.25] | **0.0066**, 0.26, [-0.01, 0.52] | 0.2514, 0.01, [-0.26, 0.28] | 0.1079, -0.21, [-0.48, 0.05] |
| 43 | SFOF R | **0.0473**, 0.3, [0.03, 0.57] | 0.1104, 0.23, [-0.03, 0.5] | 0.4766, 0.01, [-0.25, 0.28] | 0.0518, -0.29, [-0.56, -0.03] | 0.407, -0.13, [-0.4, 0.14] | 0.1882, -0.19, [-0.46, 0.07] | 0.2845, -0.15, [-0.42, 0.11] | 0.2982, -0.16, [-0.42, 0.11] | 0.4832, 0.02, [-0.24, 0.29] | 0.1892, 0.19, [-0.08, 0.46] | 0.3733, 0.08, [-0.19, 0.34] | 0.3024, -0.12, [-0.38, 0.15] |
| 44 | SFOF L | 0.1389, 0.0, [-0.26, 0.27] | 0.07, 0.02, [-0.24, 0.29] | 0.0786, 0.03, [-0.24, 0.29] | 0.456, -0.01, [-0.27, 0.26] | 0.38, -0.03, [-0.29, 0.24] | 0.3187, -0.08, [-0.35, 0.18] | 0.385, -0.04, [-0.3, 0.23] | 0.4496, 0.09, [-0.17, 0.36] | 0.3985, 0.07, [-0.2, 0.33] | 0.4353, 0.01, [-0.26, 0.27] | 0.1702, 0.1, [-0.16, 0.37] | 0.416, 0.05, [-0.21, 0.32] |
| 45 | IFOF R | **0.0208**, 0.36, [0.09, 0.63] | **0.0238**, 0.29, [0.02, 0.56] | 0.3698, 0.02, [-0.24, 0.29] | 0.0583, -0.36, [-0.63, -0.09] | 0.0628, -0.29, [-0.56, -0.02] | **0.0465**, -0.35, [-0.62, -0.08] | 0.4426, -0.06, [-0.32, 0.21] | 0.1319, -0.22, [-0.49, 0.05] | 0.1749, 0.15, [-0.12, 0.41] | 0.2158, 0.15, [-0.12, 0.42] | 0.1475, 0.18, [-0.09, 0.45] | 0.3772, -0.11, [-0.37, 0.16] |
| 46 | IFOF L | 0.1067, 0.07, [-0.2, 0.34] | 0.0817, 0.04, [-0.23, 0.3] | 0.3161, -0.03, [-0.3, 0.24] | 0.2301, -0.16, [-0.43, 0.1] | 0.4032, -0.08, [-0.34, 0.19] | 0.2614, -0.17, [-0.43, 0.1] | 0.4483, 0.07, [-0.2, 0.33] | 0.4527, 0.0, [-0.26, 0.27] | 0.1065, 0.1, [-0.17, 0.37] | 0.4616, -0.08, [-0.34, 0.19] | 0.0739, 0.1, [-0.16, 0.37] | 0.4644, -0.0, [-0.27, 0.26] |
| 47 | UF R | 0.1038, 0.22, [-0.05, 0.48] | 0.1228, 0.21, [-0.06, 0.48] | 0.2845, 0.08, [-0.19, 0.34] | 0.2261, -0.13, [-0.4, 0.13] | 0.104, -0.25, [-0.52, 0.01] | 0.1589, -0.24, [-0.51, 0.03] | 0.1706, -0.23, [-0.5, 0.04] | 0.1457, -0.23, [-0.5, 0.04] | 0.4116, -0.11, [-0.37, 0.16] | 0.0898, 0.3, [0.04, 0.57] | 0.4616, -0.03, [-0.3, 0.24] | 0.2614, -0.14, [-0.4, 0.13] |
| 48 | UF L | 0.361, -0.0, [-0.27, 0.26] | 0.3496, -0.04, [-0.31, 0.23] | 0.3583, -0.08, [-0.35, 0.19] | 0.2301, -0.11, [-0.37, 0.16] | 0.1671, -0.15, [-0.41, 0.12] | 0.1945, -0.18, [-0.44, 0.09] | 0.1044, -0.22, [-0.49, 0.05] | 0.1638, -0.18, [-0.44, 0.09] | 0.3087, -0.15, [-0.42, 0.12] | 0.4095, -0.01, [-0.28, 0.25] | 0.4816, -0.07, [-0.34, 0.19] | 0.2334, -0.13, [-0.4, 0.14] |
| 49 | TPTM R | 0.0614, 0.35, [0.08, 0.62] | 0.0518, 0.38, [0.11, 0.65] | **0.0371**, 0.4, [0.13, 0.67] | 0.2706, -0.16, [-0.43, 0.11] | 0.0583, -0.31, [-0.58, -0.04] | **0.0056**, -0.49, [-0.76, -0.22] | **0.0085**, -0.47, [-0.74, -0.2] | 0.1144, -0.23, [-0.5, 0.04] | 0.475, 0.0, [-0.26, 0.27] | 0.1341, 0.18, [-0.09, 0.44] | 0.2113, 0.13, [-0.13, 0.4] | 0.4053, -0.09, [-0.35, 0.18] |
| 50 | TPTM L | 0.1185, 0.2, [-0.07, 0.46] | 0.1273, 0.22, [-0.05, 0.48] | 0.1656, 0.23, [-0.04, 0.49] | 0.3328, -0.06, [-0.32, 0.21] | 0.2396, -0.09, [-0.36, 0.18] | 0.0746, -0.24, [-0.51, 0.03] | 0.3406, -0.09, [-0.35, 0.18] | 0.3906, -0.05, [-0.31, 0.22] | 0.4406, 0.0, [-0.26, 0.27] | 0.1185, -0.23, [-0.49, 0.04] | 0.4669, -0.06, [-0.33, 0.2] | 0.2009, -0.15, [-0.42, 0.12] |

Table S 5: Early psychosis: region of interest heatmap group comparisons. Respectively: FDR p-val, Effect size, [low-CI, high-CI]. HC: healthy controls, Y: young, O: old, EP: early psychosis, SZ: chronic schizophrenic.

| | metric | RD | MD | AD | FA | RK | MK | AK | $f$ | $D_a$ | $D_{e,\parallel}$ | $D_{e,\perp}$ | $c_2$ |
|---|---|---|---|---|---|---|---|---|---|---|---|---|---|
| 1 | MIDCP | 0.2468, 0.2, [-0.16, 0.55] | 0.1631, 0.21, [-0.15, 0.57] | 0.1195, 0.23, [-0.13, 0.59] | 0.474, -0.09, [-0.45, 0.27] | **0.0378**, -0.49, [-0.86, -0.13] | 0.0716, -0.38, [-0.74, -0.02] | 0.2017, -0.2, [-0.56, 0.16] | 0.1882, -0.32, [-0.68, 0.04] | 0.3273, -0.01, [-0.37, 0.35] | 0.1684, 0.13, [-0.23, 0.49] | 0.2818, 0.04, [-0.32, 0.39] | 0.2646, -0.06, [-0.42, 0.3] |
| 2 | PCT | 0.2342, -0.21, [-0.57, 0.15] | 0.1871, -0.27, [-0.62, 0.09] | 0.2301, -0.28, [-0.64, 0.08] | 0.4203, 0.01, [-0.35, 0.37] | 0.1251, 0.23, [-0.13, 0.59] | 0.1183, 0.22, [-0.13, 0.58] | 0.221, 0.22, [-0.13, 0.58] | 0.2896, 0.05, [-0.31, 0.41] | 0.2261, -0.26, [-0.62, 0.1] | 0.0755, -0.36, [-0.72, 0.0] | 0.3951, -0.12, [-0.48, 0.24] | 0.3916, -0.11, [-0.47, 0.25] |
| 3 | GCC | 0.0866, 0.35, [-0.01, 0.71] | **0.0407**, 0.45, [0.09, 0.81] | 0.0716, 0.44, [0.08, 0.8] | 0.2301, -0.19, [-0.54, 0.17] | 0.1991, -0.24, [-0.6, 0.12] | 0.104, -0.28, [-0.64, 0.08] | 0.4008, 0.07, [-0.28, 0.43] | 0.3558, -0.02, [-0.37, 0.34] | **0.0133**, 0.62, [0.25, 0.98] | 0.1228, 0.27, [-0.09, 0.63] | 0.104, 0.38, [0.02, 0.74] | 0.4264, -0.08, [-0.44, 0.27] |
| 4 | BCC | 0.1228, 0.33, [-0.03, 0.68] | 0.0745, 0.38, [0.02, 0.74] | 0.094, 0.34, [-0.02, 0.7] | 0.1901, -0.25, [-0.61, 0.11] | 0.1553, -0.34, [-0.7, 0.02] | 0.0563, -0.48, [-0.84, -0.11] | **0.0218**, -0.49, [-0.85, -0.13] | 0.1103, -0.37, [-0.73, -0.01] | 0.1134, 0.3, [-0.06, 0.65] | **0.0249**, 0.45, [0.09, 0.81] | 0.1753, 0.23, [-0.13, 0.59] | 0.2401, -0.2, [-0.56, 0.15] |
| 5 | SCC | 0.2147, 0.33, [-0.03, 0.69] | 0.149, 0.35, [-0.01, 0.71] | 0.139, 0.29, [-0.07, 0.65] | 0.3544, -0.22, [-0.58, 0.14] | 0.487, -0.11, [-0.47, 0.24] | 0.1319, -0.37, [-0.73, -0.01] | 0.2894, -0.22, [-0.58, 0.14] | 0.3124, 0.06, [-0.29, 0.42] | 0.2847, 0.1, [-0.25, 0.46] | 0.2638, -0.15, [-0.51, 0.2] | 0.0981, 0.36, [0.0, 0.72] | 0.4557, 0.01, [-0.35, 0.36] |
| 6 | FX | 0.1061, 0.34, [-0.02, 0.69] | 0.0941, 0.34, [-0.02, 0.7] | 0.102, 0.31, [-0.05, 0.66] | 0.1596, -0.28, [-0.63, 0.08] | **0.0489**, -0.42, [-0.78, -0.06] | **0.0367**, -0.48, [-0.84, -0.12] | 0.1286, -0.31, [-0.67, 0.05] | 0.0628, -0.41, [-0.77, -0.05] | 0.1183, -0.35, [-0.71, 0.01] | 0.1315, -0.35, [-0.71, 0.01] | 0.3543, -0.17, [-0.53, 0.19] | 0.2901, -0.28, [-0.64, 0.08] |
| 7 | CST R | 0.4695, 0.01, [-0.35, 0.36] | 0.4511, 0.01, [-0.35, 0.36] | 0.3735, 0.01, [-0.34, 0.37] | 0.487, 0.01, [-0.35, 0.37] | 0.4277, -0.02, [-0.38, 0.34] | 0.3544, 0.05, [-0.3, 0.41] | 0.3149, 0.14, [-0.22, 0.5] | 0.3798, -0.03, [-0.39, 0.33] | 0.2034, 0.04, [-0.32, 0.4] | 0.2518, -0.18, [-0.54, 0.18] | 0.4234, -0.01, [-0.37, 0.35] | 0.4008, -0.06, [-0.42, 0.3] |
| 8 | CST L | 0.3955, -0.05, [-0.4, 0.31] | 0.487, 0.03, [-0.33, 0.38] | 0.3289, 0.13, [-0.23, 0.49] | 0.3479, 0.09, [-0.27, 0.44] | 0.475, -0.09, [-0.44, 0.27] | 0.4234, -0.17, [-0.52, 0.19] | 0.4616, 0.08, [-0.28, 0.43] | 0.4578, 0.0, [-0.35, 0.36] | 0.0551, 0.28, [-0.08, 0.64] | 0.2439, -0.23, [-0.59, 0.13] | 0.2982, -0.08, [-0.44, 0.28] | 0.0743, 0.07, [-0.28, 0.43] |
| 9 | ML R | 0.0598, 0.26, [-0.1, 0.63] | 0.0775, 0.2, [-0.16, 0.56] | 0.3754, 0.05, [-0.31, 0.41] | 0.0662, -0.24, [-0.61, 0.12] | 0.1299, -0.28, [-0.64, 0.08] | 0.3298, -0.14, [-0.51, 0.22] | 0.474, 0.08, [-0.28, 0.44] | 0.1206, -0.24, [-0.6, 0.12] | 0.2091, 0.15, [-0.21, 0.51] | 0.1499, 0.15, [-0.22, 0.51] | 0.2463, 0.14, [-0.22, 0.5] | 0.2062, -0.23, [-0.59, 0.13] |
| 10 | ML L | 0.1758, 0.13, [-0.23, 0.49] | 0.2856, 0.01, [-0.35, 0.37] | 0.3599, -0.13, [-0.49, 0.23] | 0.1945, -0.19, [-0.55, 0.17] | 0.3361, -0.11, [-0.47, 0.25] | 0.4283, -0.12, [-0.48, 0.24] | 0.2851, 0.19, [-0.17, 0.55] | 0.2896, -0.09, [-0.45, 0.27] | 0.3805, 0.0, [-0.36, 0.36] | 0.4008, -0.01, [-0.37, 0.35] | 0.3423, 0.05, [-0.31, 0.41] | 0.3112, -0.12, [-0.48, 0.24] |
| 11 | ICP R | 0.3044, 0.16, [-0.21, 0.53] | 0.379, 0.06, [-0.31, 0.42] | 0.2647, -0.11, [-0.47, 0.26] | 0.2951, -0.2, [-0.57, 0.17] | 0.1186, -0.25, [-0.62, 0.12] | 0.1962, -0.21, [-0.58, 0.16] | 0.2261, 0.24, [-0.13, 0.61] | 0.2301, -0.16, [-0.52, 0.21] | 0.4116, 0.12, [-0.25, 0.49] | 0.487, 0.01, [-0.35, 0.38] | 0.4616, 0.06, [-0.3, 0.43] | 0.3271, -0.17, [-0.54, 0.2] |
| 12 | ICP L | 0.4116, -0.06, [-0.43, 0.31] | 0.4979, -0.14, [-0.5, 0.23] | 0.2946, -0.22, [-0.58, 0.15] | 0.3922, -0.14, [-0.5, 0.23] | 0.2437, -0.21, [-0.58, 0.16] | 0.2975, -0.17, [-0.54, 0.2] | 0.2226, 0.2, [-0.17, 0.57] | 0.2418, -0.25, [-0.62, 0.12] | 0.4806, 0.0, [-0.37, 0.37] | 0.474, -0.03, [-0.4, 0.34] | 0.3482, -0.11, [-0.48, 0.26] | 0.4669, -0.01, [-0.38, 0.36] |
| 13 | SCP R | **0.0238**, 0.53, [0.17, 0.9] | **0.0327**, 0.49, [0.13, 0.86] | 0.0819, 0.32, [-0.04, 0.68] | **0.0114**, -0.39, [-0.75, -0.03] | 0.473, -0.1, [-0.46, 0.25] | 0.2379, -0.17, [-0.53, 0.18] | 0.4116, 0.12, [-0.24, 0.48] | 0.3187, 0.1, [-0.26, 0.46] | 0.2367, -0.08, [-0.44, 0.27] | 0.3194, 0.16, [-0.2, 0.52] | **0.015**, 0.56, [0.2, 0.92] | 0.1551, -0.15, [-0.51, 0.21] |
| 14 | SCP L | 0.0533, 0.36, [-0.0, 0.72] | 0.057, 0.37, [0.01, 0.73] | 0.1075, 0.3, [-0.06, 0.66] | 0.1431, -0.24, [-0.6, 0.12] | 0.4203, -0.01, [-0.37, 0.35] | 0.2301, -0.19, [-0.55, 0.16] | 0.3542, 0.16, [-0.2, 0.51] | 0.1724, 0.28, [-0.08, 0.64] | 0.4506, 0.06, [-0.3, 0.42] | 0.3298, 0.05, [-0.31, 0.41] | **0.0088**, 0.63, [0.26, 0.99] | 0.3372, 0.15, [-0.2, 0.51] |
| 15 | CP R | 0.3919, 0.2, [-0.16, 0.56] | 0.4669, 0.15, [-0.21, 0.51] | 0.4849, 0.06, [-0.3, 0.41] | 0.2262, -0.2, [-0.55, 0.16] | 0.3194, 0.06, [-0.3, 0.42] | 0.1882, 0.09, [-0.27, 0.45] | 0.0869, 0.36, [-0.0, 0.72] | 0.3185, 0.09, [-0.26, 0.45] | 0.0746, 0.34, [-0.02, 0.7] | 0.2301, -0.22, [-0.57, 0.14] | 0.143, 0.36, [-0.0, 0.72] | 0.4583, -0.06, [-0.41, 0.3] |
| 16 | CP L | 0.4075, 0.16, [-0.2, 0.51] | 0.4862, 0.03, [-0.32, 0.39] | 0.1882, -0.17, [-0.52, 0.19] | 0.1822, -0.29, [-0.65, 0.06] | 0.3968, -0.19, [-0.54, 0.17] | 0.198, 0.05, [-0.31, 0.4] | 0.1144, 0.31, [-0.05, 0.67] | 0.1533, -0.32, [-0.68, 0.04] | 0.4585, 0.02, [-0.34, 0.37] | 0.3745, -0.02, [-0.38, 0.34] | 0.3662, -0.06, [-0.42, 0.29] | 0.1253, -0.34, [-0.7, 0.02] |
| 17 | ALIC R | 0.1351, 0.39, [0.02, 0.74] | 0.214, 0.29, [-0.07, 0.65] | 0.4557, 0.03, [-0.33, 0.39] | 0.1195, -0.35, [-0.71, 0.01] | 0.0895, -0.31, [-0.67, 0.05] | 0.1734, -0.18, [-0.54, 0.18] | 0.4352, 0.0, [-0.36, 0.36] | 0.118, -0.27, [-0.63, 0.08] | 0.2981, 0.07, [-0.28, 0.43] | 0.2017, 0.24, [-0.12, 0.59] | 0.3919, 0.12, [-0.23, 0.48] | 0.1702, -0.28, [-0.64, 0.08] |
| 18 | ALIC L | 0.2229, 0.25, [-0.11, 0.61] | 0.1945, 0.2, [-0.15, 0.56] | 0.3125, 0.08, [-0.28, 0.44] | 0.2852, -0.17, [-0.53, 0.18] | **0.0047**, -0.63, [-0.99, -0.26] | 0.2463, -0.2, [-0.55, 0.16] | 0.214, -0.17, [-0.53, 0.19] | **0.0047**, -0.59, [-0.95, -0.22] | 0.2463, 0.21, [-0.15, 0.56] | **0.0041**, 0.68, [0.31, 1.04] | 0.126, -0.3, [-0.66, 0.06] | 0.1596, -0.29, [-0.65, 0.07] |
| 19 | PLIC R | 0.2894, 0.22, [-0.14, 0.58] | 0.1013, 0.33, [-0.03, 0.69] | 0.2039, 0.29, [-0.07, 0.65] | 0.474, -0.01, [-0.37, 0.35] | 0.4318, 0.03, [-0.33, 0.38] | 0.4881, -0.08, [-0.44, 0.27] | 0.2113, 0.18, [-0.18, 0.53] | 0.352, 0.08, [-0.28, 0.44] | 0.0571, 0.41, [0.05, 0.77] | 0.3922, -0.09, [-0.45, 0.26] | 0.2119, 0.3, [-0.06, 0.65] | 0.2547, 0.19, [-0.17, 0.55] |
| 20 | PLIC L | 0.4557, 0.03, [-0.33, 0.38] | 0.3968, 0.11, [-0.25, 0.47] | 0.2845, 0.22, [-0.13, 0.58] | 0.4313, 0.04, [-0.32, 0.39] | 0.4008, -0.2, [-0.56, 0.16] | 0.3804, -0.25, [-0.61, 0.11] | 0.2463, -0.2, [-0.55, 0.16] | 0.2917, -0.25, [-0.61, 0.11] | 0.2861, 0.26, [-0.1, 0.62] | 0.0746, 0.32, [-0.04, 0.68] | 0.3639, -0.08, [-0.44, 0.28] | 0.3427, 0.14, [-0.22, 0.49] |
| 21 | RPIC R | 0.2191, 0.26, [-0.1, 0.62] | 0.0862, 0.27, [-0.08, 0.63] | 0.1509, 0.25, [-0.11, 0.6] | 0.3553, -0.21, [-0.57, 0.14] | 0.2982, -0.17, [-0.53, 0.19] | 0.1631, -0.34, [-0.69, 0.02] | 0.4496, 0.04, [-0.32, 0.39] | 0.3159, -0.07, [-0.43, 0.29] | 0.0982, 0.31, [-0.05, 0.67] | 0.4207, 0.07, [-0.29, 0.42] | 0.1684, 0.29, [-0.07, 0.65] | 0.3557, 0.11, [-0.24, 0.47] |
| 22 | RPIC L | 0.0766, 0.25, [-0.11, 0.61] | 0.0536, 0.27, [-0.09, 0.63] | 0.0949, 0.25, [-0.11, 0.61] | 0.2437, -0.19, [-0.55, 0.17] | 0.2148, -0.2, [-0.56, 0.15] | 0.2357, -0.22, [-0.58, 0.14] | 0.3747, -0.11, [-0.47, 0.24] | 0.2103, -0.24, [-0.6, 0.12] | 0.2813, 0.14, [-0.22, 0.5] | 0.1195, 0.33, [-0.03, 0.69] | 0.3353, 0.14, [-0.22, 0.49] | 0.4682, -0.08, [-0.43, 0.28] |
| 23 | ACR R | **0.0102**, 0.63, [0.27, 1.0] | **0.0073**, 0.69, [0.32, 1.05] | **0.037**, 0.48, [0.12, 0.84] | 0.0662, -0.41, [-0.77, -0.05] | 0.0846, -0.38, [-0.74, -0.02] | 0.0583, -0.44, [-0.8, -0.08] | 0.0746, -0.43, [-0.79, -0.07] | 0.0581, -0.34, [-0.7, 0.02] | 0.2646, 0.26, [-0.1, 0.62] | **0.0041**, 0.65, [0.28, 1.01] | 0.0895, 0.34, [-0.02, 0.7] | 0.0804, -0.43, [-0.79, -0.07] |
| 24 | ACR L | **0.0379**, 0.48, [0.11, 0.84] | **0.0378**, 0.48, [0.12, 0.84] | 0.1471, 0.33, [-0.03, 0.68] | 0.1056, -0.31, [-0.67, 0.05] | **0.0114**, -0.57, [-0.93, -0.2] | **0.0228**, -0.55, [-0.91, -0.19] | **0.0355**, -0.43, [-0.79, -0.06] | **0.0181**, -0.48, [-0.84, -0.11] | 0.1882, 0.2, [-0.16, 0.55] | **0.0349**, 0.5, [0.14, 0.87] | 0.2891, 0.25, [-0.11, 0.61] | 0.174, -0.24, [-0.6, 0.12] |
| 25 | SCR R | **0.0342**, 0.43, [0.07, 0.79] | 0.0669, 0.37, [0.01, 0.73] | 0.3423, 0.17, [-0.18, 0.53] | 0.1631, -0.26, [-0.62, 0.1] | **0.037**, -0.5, [-0.86, -0.14] | **0.0093**, -0.6, [-0.96, -0.23] | 0.0572, -0.46, [-0.82, -0.1] | 0.1794, -0.33, [-0.69, 0.03] | 0.0542, 0.42, [0.06, 0.78] | 0.22, 0.33, [-0.03, 0.69] | 0.479, 0.03, [-0.33, 0.38] | 0.3638, -0.09, [-0.45, 0.26] |
| 26 | SCR L | 0.1684, 0.25, [-0.11, 0.61] | 0.1134, 0.3, [-0.06, 0.65] | 0.1157, 0.3, [-0.06, 0.66] | 0.2301, -0.11, [-0.47, 0.24] | **0.0048**, -0.67, [-1.04, -0.3] | **0.0023**, -0.7, [-1.07, -0.34] | **0.0001**, -0.84, [-1.21, -0.47] | **0.0041**, -0.69, [-1.05, -0.32] | 0.174, 0.27, [-0.08, 0.63] | **0.0025**, 0.78, [0.41, 1.15] | 0.1058, -0.28, [-0.63, 0.08] | 0.3292, -0.06, [-0.42, 0.29] |
| 27 | PCR R | 0.0669, 0.37, [0.01, 0.73] | **0.0286**, 0.45, [0.09, 0.81] | **0.0431**, 0.44, [0.08, 0.8] | 0.2982, -0.12, [-0.48, 0.23] | 0.197, -0.29, [-0.65, 0.07] | 0.0658, -0.42, [-0.78, -0.06] | 0.058, -0.4, [-0.76, -0.04] | 0.3539, -0.2, [-0.55, 0.16] | 0.1065, 0.39, [0.03, 0.75] | 0.0981, 0.28, [-0.08, 0.63] | 0.2023, 0.26, [-0.1, 0.61] | 0.4582, 0.02, [-0.34, 0.38] |
| 28 | PCR L | 0.2331, 0.18, [-0.18, 0.54] | 0.1314, 0.2, [-0.16, 0.56] | 0.1513, 0.21, [-0.14, 0.57] | 0.4353, -0.02, [-0.38, 0.33] | 0.1586, -0.29, [-0.65, 0.07] | **0.0152**, -0.5, [-0.86, -0.13] | 0.0887, -0.37, [-0.73, -0.01] | 0.4824, -0.07, [-0.42, 0.29] | 0.057, 0.4, [0.04, 0.76] | 0.4915, 0.08, [-0.28, 0.44] | 0.126, 0.26, [-0.1, 0.61] | 0.2272, 0.17, [-0.19, 0.52] |
| 29 | PTR R | 0.1044, 0.33, [-0.03, 0.69] | **0.0378**, 0.37, [0.01, 0.73] | **0.0363**, 0.3, [-0.06, 0.66] | 0.3344, -0.16, [-0.51, 0.2] | 0.1596, -0.38, [-0.74, -0.02] | 0.0551, -0.53, [-0.89, -0.17] | 0.0518, -0.45, [-0.81, -0.09] | 0.2206, -0.34, [-0.7, 0.02] | 0.1571, 0.2, [-0.16, 0.55] | 0.1285, 0.29, [-0.07, 0.65] | 0.1189, 0.31, [-0.05, 0.66] | 0.4133, 0.11, [-0.25, 0.47] |
| 30 | PTR L | **0.0378**, 0.47, [0.1, 0.83] | **0.0378**, 0.45, [0.08, 0.81] | 0.1611, 0.28, [-0.07, 0.64] | 0.0772, -0.41, [-0.77, -0.05] | **0.0228**, -0.5, [-0.86, -0.13] | **0.0085**, -0.56, [-0.92, -0.2] | 0.0662, -0.38, [-0.74, -0.02] | 0.0928, -0.33, [-0.69, 0.03] | 0.2421, 0.13, [-0.23, 0.49] | 0.0518, 0.4, [0.04, 0.76] | 0.107, 0.4, [0.04, 0.76] | 0.2975, -0.19, [-0.55, 0.16] |
| 31 | SAGSTR R | 0.0536, 0.46, [0.09, 0.82] | 0.057, 0.42, [0.06, 0.78] | 0.2009, 0.23, [-0.13, 0.59] | 0.1056, -0.26, [-0.62, 0.1] | **0.0181**, -0.53, [-0.89, -0.16] | **0.0092**, -0.65, [-1.01, -0.28] | 0.0866, -0.36, [-0.72, -0.0] | 0.0595, -0.31, [-0.67, 0.05] | 0.1079, 0.31, [-0.05, 0.66] | 0.0866, 0.28, [-0.08, 0.64] | 0.2062, 0.29, [-0.07, 0.64] | 0.2653, -0.05, [-0.41, 0.31] |
| 32 | SAGSTR L | 0.0895, 0.3, [-0.06, 0.66] | 0.0775, 0.28, [-0.08, 0.64] | 0.2285, 0.17, [-0.19, 0.53] | 0.139, -0.3, [-0.65, 0.06] | **0.037**, -0.42, [-0.78, -0.06] | **0.0485**, -0.43, [-0.79, -0.06] | 0.1628, -0.31, [-0.67, 0.05] | **0.0365**, -0.4, [-0.76, -0.03] | 0.3303, 0.04, [-0.32, 0.39] | 0.1, 0.3, [-0.06, 0.66] | 0.3552, 0.11, [-0.24, 0.47] | 0.2813, -0.16, [-0.52, 0.2] |
| 33 | EC R | 0.4353, 0.09, [-0.27, 0.44] | 0.2982, 0.14, [-0.22, 0.49] | 0.229, 0.19, [-0.17, 0.54] | 0.4512, 0.11, [-0.25, 0.47] | 0.1631, -0.26, [-0.62, 0.1] | 0.1562, -0.25, [-0.6, 0.11] | 0.485, -0.03, [-0.39, 0.32] | 0.2489, -0.2, [-0.56, 0.15] | 0.2813, 0.24, [-0.12, 0.6] | 0.4358, 0.08, [-0.28, 0.43] | 0.4834, 0.02, [-0.33, 0.38] | 0.2547, 0.22, [-0.14, 0.58] |
| 34 | EC L | 0.4188, 0.09, [-0.26, 0.45] | 0.3201, 0.08, [-0.27, 0.44] | 0.4176, 0.06, [-0.3, 0.42] | 0.4773, 0.01, [-0.35, 0.36] | 0.4282, -0.01, [-0.37, 0.35] | 0.397, -0.05, [-0.41, 0.31] | 0.2017, 0.26, [-0.1, 0.62] | 0.2229, 0.11, [-0.24, 0.47] | 0.0945, 0.36, [-0.0, 0.72] | 0.3273, -0.06, [-0.42, 0.29] | 0.2301, 0.16, [-0.2, 0.52] | 0.2396, 0.17, [-0.19, 0.53] |
| 35 | CING/CG R | 0.1148, 0.29, [-0.06, 0.65] | 0.1875, 0.23, [-0.13, 0.59] | 0.4858, -0.02, [-0.37, 0.34] | 0.2272, -0.26, [-0.62, 0.1] | 0.1476, -0.31, [-0.67, 0.05] | 0.1389, -0.36, [-0.71, 0.0] | 0.1471, -0.23, [-0.59, 0.13] | 0.2075, -0.26, [-0.62, 0.1] | 0.2405, 0.17, [-0.18, 0.53] | 0.19, 0.25, [-0.1, 0.61] | 0.4557, 0.05, [-0.3, 0.41] | 0.4364, -0.13, [-0.48, 0.23] |
| 36 | CING/CG L | 0.0804, 0.39, [0.02, 0.74] | 0.0684, 0.35, [-0.01, 0.71] | 0.4487, 0.09, [-0.27, 0.44] | 0.1684, -0.29, [-0.65, 0.07] | 0.0898, -0.37, [-0.73, -0.01] | 0.1143, -0.42, [-0.78, -0.06] | 0.1389, -0.3, [-0.66, 0.06] | 0.104, -0.33, [-0.69, 0.03] | 0.4008, 0.12, [-0.24, 0.48] | 0.0615, 0.36, [-0.0, 0.72] | 0.2374, 0.19, [-0.17, 0.55] | 0.4008, -0.16, [-0.52, 0.19] |
| 37 | CING/HIP R | 0.0895, 0.53, [0.17, 0.89] | 0.0723, 0.51, [0.15, 0.87] | **0.0471**, 0.42, [0.05, 0.78] | 0.1157, -0.29, [-0.64, 0.07] | 0.38, 0.17, [-0.19, 0.52] | 0.4164, 0.05, [-0.31, 0.41] | 0.2285, 0.19, [-0.17, 0.55] | 0.428, 0.13, [-0.23, 0.48] | 0.2113, 0.19, [-0.17, 0.54] | 0.2901, 0.12, [-0.23, 0.48] | **0.0344**, 0.57, [0.21, 0.94] | 0.2553, -0.13, [-0.49, 0.23] |
| 38 | CING/HIP L | 0.1553, 0.37, [0.01, 0.73] | 0.0786, 0.38, [0.02, 0.74] | 0.0823, 0.34, [-0.02, 0.7] | 0.3951, -0.09, [-0.44, 0.27] | 0.2896, 0.15, [-0.21, 0.51] | 0.4557, -0.01, [-0.36, 0.35] | 0.3148, 0.15, [-0.2, 0.51] | 0.3552, 0.2, [-0.16, 0.56] | **0.0378**, 0.41, [0.05, 0.77] | 0.4164, -0.11, [-0.47, 0.24] | 0.104, 0.39, [0.03, 0.75] | 0.4506, -0.06, [-0.42, 0.29] |
| 39 | FX/STRIA R | 0.0945, 0.43, [0.07, 0.79] | 0.082, 0.43, [0.07, 0.79] | 0.1098, 0.41, [0.05, 0.77] | 0.3024, -0.16, [-0.52, 0.2] | 0.1341, -0.3, [-0.66, 0.06] | 0.1413, -0.29, [-0.65, 0.07] | 0.3353, -0.16, [-0.51, 0.2] | 0.1659, -0.34, [-0.7, 0.02] | 0.3161, -0.15, [-0.51, 0.2] | 0.1295, 0.34, [-0.02, 0.7] | 0.1885, 0.28, [-0.08, 0.63] | 0.3922, -0.11, [-0.47, 0.25] |
| 40 | FX/STRIA L | 0.104, 0.44, [0.07, 0.8] | 0.104, 0.39, [0.03, 0.75] | 0.1534, 0.27, [-0.09, 0.63] | 0.174, -0.29, [-0.65, 0.07] | 0.2896, -0.15, [-0.51, 0.21] | 0.2659, -0.1, [-0.46, 0.26] | 0.474, -0.03, [-0.39, 0.32] | 0.304, -0.22, [-0.58, 0.14] | 0.3867, -0.12, [-0.47, 0.24] | 0.2017, 0.25, [-0.11, 0.61] | 0.1513, 0.29, [-0.07, 0.65] | 0.2017, -0.32, [-0.68, 0.04] |
| 41 | SLF R | 0.0613, 0.35, [-0.01, 0.71] | **0.0365**, 0.44, [0.08, 0.8] | 0.0743, 0.42, [0.06, 0.78] | 0.2317, -0.17, [-0.53, 0.18] | 0.0631, -0.37, [-0.73, -0.01] | **0.0336**, -0.47, [-0.83, -0.11] | **0.0181**, -0.56, [-0.93, -0.2] | 0.1289, -0.3, [-0.66, 0.06] | 0.1183, 0.33, [-0.03, 0.69] | **0.0428**, 0.38, [0.02, 0.74] | 0.3583, 0.13, [-0.22, 0.49] | 0.4834, -0.03, [-0.39, 0.32] |
| 42 | SLF L | 0.1684, 0.3, [-0.06, 0.66] | 0.0739, 0.36, [-0.0, 0.72] | 0.0851, 0.38, [0.02, 0.74] | 0.4557, -0.1, [-0.46, 0.25] | **0.014**, -0.48, [-0.84, -0.12] | **0.007**, -0.6, [-0.97, -0.24] | **0.0054**, -0.61, [-0.97, -0.24] | **0.0267**, -0.42, [-0.78, -0.05] | 0.197, 0.32, [-0.04, 0.67] | **0.0202**, 0.4, [0.04, 0.76] | 0.4054, -0.0, [-0.36, 0.36] | 0.4862, -0.01, [-0.37, 0.35] |
| 43 | SFOF R | **0.014**, 0.57, [0.21, 0.93] | 0.083, 0.44, [0.07, 0.8] | 0.4721, 0.04, [-0.32, 0.39] | **0.0295**, -0.49, [-0.85, -0.13] | 0.2357, -0.26, [-0.62, 0.1] | 0.0858, -0.32, [-0.68, 0.04] | 0.3344, 0.08, [-0.28, 0.44] | 0.4037, -0.07, [-0.43, 0.28] | 0.2301, 0.17, [-0.19, 0.53] | 0.4849, 0.03, [-0.33, 0.39] | 0.0957, 0.33, [-0.03, 0.69] | 0.2301, -0.22, [-0.57, 0.14] |
| 44 | SFOF L | 0.1164, 0.41, [0.05, 0.77] | 0.1677, 0.33, [-0.03, 0.69] | 0.2271, 0.19, [-0.17, 0.55] | 0.2343, -0.26, [-0.61, 0.1] | 0.0684, -0.34, [-0.7, 0.02] | **0.007**, -0.61, [-0.97, -0.24] | 0.0818, -0.42, [-0.78, -0.06] | 0.2396, -0.13, [-0.49, 0.23] | 0.4824, 0.05, [-0.3, 0.41] | 0.3055, 0.18, [-0.17, 0.54] | 0.1889, 0.22, [-0.13, 0.58] | 0.3336, -0.14, [-0.49, 0.22] |
| 45 | IFOF R | 0.3093, 0.11, [-0.25, 0.47] | 0.2206, 0.2, [-0.16, 0.55] | 0.1645, 0.27, [-0.09, 0.62] | 0.4674, 0.04, [-0.31, 0.4] | 0.2357, -0.13, [-0.49, 0.23] | 0.402, -0.08, [-0.44, 0.27] | 0.474, -0.08, [-0.44, 0.27] | 0.188, -0.15, [-0.5, 0.21] | 0.2075, 0.25, [-0.11, 0.61] | 0.3044, 0.18, [-0.18, 0.54] | 0.4832, -0.03, [-0.38, 0.33] | 0.4757, -0.0, [-0.36, 0.35] |
| 46 | IFOF L | 0.4177, 0.08, [-0.28, 0.44] | 0.3336, 0.14, [-0.22, 0.49] | 0.1769, 0.2, [-0.16, 0.56] | 0.4454, 0.06, [-0.29, 0.42] | 0.4645, 0.01, [-0.35, 0.37] | 0.4557, 0.03, [-0.33, 0.39] | 0.3344, 0.05, [-0.3, 0.41] | 0.475, -0.0, [-0.36, 0.35] | 0.148, 0.26, [-0.09, 0.62] | 0.4834, 0.08, [-0.27, 0.44] | 0.4264, 0.04, [-0.31, 0.4] | 0.4616, 0.01, [-0.34, 0.37] |
| 47 | UF R | 0.2851, 0.1, [-0.26, 0.46] | 0.1659, 0.25, [-0.11, 0.61] | 0.0802, 0.34, [-0.02, 0.69] | 0.4285, 0.08, [-0.27, 0.44] | 0.1817, -0.22, [-0.58, 0.13] | 0.4834, -0.0, [-0.36, 0.36] | 0.3624, -0.17, [-0.53, 0.19] | 0.1696, -0.26, [-0.61, 0.1] | 0.405, -0.13, [-0.48, 0.23] | **0.0218**, 0.48, [0.12, 0.84] | 0.3427, -0.14, [-0.5, 0.21] | 0.3124, -0.11, [-0.47, 0.24] |
| 48 | UF L | 0.2537, 0.18, [-0.18, 0.54] | 0.2401, 0.22, [-0.14, 0.58] | 0.1871, 0.19, [-0.17, 0.54] | 0.4557, 0.01, [-0.35, 0.37] | 0.4449, 0.0, [-0.35, 0.36] | 0.4563, -0.02, [-0.38, 0.34] | 0.4506, 0.05, [-0.31, 0.41] | 0.4207, -0.02, [-0.38, 0.33] | 0.3259, 0.22, [-0.14, 0.57] | 0.4531, 0.02, [-0.33, 0.38] | 0.2349, 0.23, [-0.13, 0.59] | 0.4797, 0.0, [-0.35, 0.36] |
| 49 | TPTM R | 0.1164, 0.3, [-0.06, 0.66] | 0.0982, 0.36, [-0.0, 0.72] | 0.0646, 0.41, [0.04, 0.76] | 0.2139, -0.14, [-0.5, 0.21] | 0.0532, -0.47, [-0.83, -0.11] | **0.016**, -0.56, [-0.93, -0.2] | **0.0164**, -0.49, [-0.85, -0.13] | 0.214, -0.2, [-0.56, 0.16] | 0.4693, -0.01, [-0.36, 0.35] | 0.1457, 0.3, [-0.06, 0.66] | 0.083, 0.35, [-0.01, 0.71] | 0.4616, 0.05, [-0.31, 0.4] |
| 50 | TPTM L | 0.1295, 0.33, [-0.03, 0.69] | 0.102, 0.34, [-0.02, 0.7] | 0.097, 0.31, [-0.05, 0.67] | 0.307, -0.15, [-0.51, 0.2] | 0.174, -0.25, [-0.61, 0.11] | 0.0863, -0.38, [-0.74, -0.02] | 0.1229, -0.28, [-0.64, 0.08] | 0.2454, -0.19, [-0.55, 0.17] | 0.251, -0.23, [-0.58, 0.13] | 0.245, -0.24, [-0.6, 0.12] | 0.245, -0.17, [-0.53, 0.19] | 0.2075, -0.27, [-0.63, 0.09] |

Table S 6: Chronic schizophrenia: region of interest heatmap group comparisons. Respectively: FDR p-val, Effect size, [low-CI, high-CI]. HC: healthy controls, Y: young, O: old, EP: early psychosis, SZ: chronic schizophrenic.
